# Serotype skewing and immune imprinting shape response to the tetravalent dengue virus Qdenga vaccine

**DOI:** 10.64898/2026.06.15.26355542

**Authors:** Luciana Conde, Valter Monteiro, Joao CR Freitas, Jameson Crandell, Lauren Lawres, Fernanda Tana, Mallery I. Breban, Jordan Polster, Fahima Akther, Abbey Porzucek, Tauyne M Ramiro, Ana B de Oliveira, Mauricio L Nogueira, Nathan D Grubaugh, David R Martinez, Pedro RJ Almeida, Daniela Weiskopf, Mauro M Teixeira, Carolina Lucas

## Abstract

Dengue remains a major global health threat, with over 390 million annual infections. The development of safe, effective, and broadly protective vaccines has been hindered by the need for tetravalent coverage and the risk of antibody-dependent enhancement (ADE). Qdenga (TAK-003) is the only dengue vaccine currently widely available globally, yet its immunological profile remains incompletely defined. Here, we characterized humoral and cellular responses, assessing the impact of the vaccine backbone and immune imprinting on vaccine-induced immunity. We conducted a longitudinal immunological study in 110 adults from a dengue-endemic region, stratified by baseline DENV serostatus, sex, and age, including older adults (≥65 years), a population excluded from efficacy trials. Plasma and PBMCs were collected before and up to five months after Qdenga vaccination to assess neutralizing antibody titers, B cell dynamics, and virus-specific T cell responses. Although Qdenga elicited B and T cell activation and memory formation across groups, our findings revealed serostatus-dependent and serotype-skewed humoral immunity. Only 8% of DENV-naïve individuals developed tetravalent responses, while one-third responded to a single serotype, almost exclusively DENV-2, the vaccine backbone. In contrast, DENV-previously exposed individuals mounted broader responses shaped by baseline serotype-specific immunity, yet neutralization against DENV-4 remained uniformly poor. Neutralizing antibody titers plateaued after the first vaccination, with no substantial increase following the second dose. These findings clarify why balanced tetravalent immunity is rarely achieved with Qdenga and demonstrate that both immune imprinting and vaccine backbone skewing limit the breadth and magnitude of the responses. This has important implications for deployment strategies, long-term protection in DENV-naïve populations, and the need for booster strategies focusing on tetravalent coverage, especially for DENV-4.

## Introduction

Dengue is the most prevalent mosquito-borne disease globally, placing nearly half of the world’s population at risk, with approximately 390 million infections annually^1^. In recent years, global dengue incidence and geographic spread have markedly increased, with the Americas reporting a record 12.6 million cases in 2024, nearly triple the previous year’s number, highlighting the urgent need for effective vaccination strategies^2^. Despite decades of research, the development of an effective dengue vaccine remains challenging due to several immunological complexities. Dengue vaccine candidates must induce balanced and durable immune responses against all four antigenically distinct dengue virus (DENV) serotypes. This is important since an imbalanced, suboptimal response can lead to antibody-dependent enhancement (ADE), particularly in individuals without prior DENV exposure, potentially exacerbating disease severity upon subsequent infections, post vaccination^3–5^. Additionally, the lack of defined immune correlates of protection complicates the assessment of vaccine efficacy and safety.

The first licensed dengue vaccine, Dengvaxia (Sanofi Pasteur), raised important safety concerns after demonstrating increased risk of severe disease in children without prior DENV exposure^6^. Its limited efficacy and safety profile highlighted critical gaps in our understanding of vaccine-induced immunity, particularly regarding immune imprinting (the shaping of subsequent immune responses by prior exposures). Although growing evidence suggests that imprinting can shape and impact vaccine effectiveness, it remains largely overlooked in vaccine design^7–9^. TAK-003 (Qdenga, Takeda), the only dengue vaccine currently widely available without pre-vaccination restrictions, is a live-attenuated tetravalent vaccine engineered using a DENV-2 backbone that expresses the prM and envelope (E) proteins of all four DENV serotypes^10^. Administered in two doses with a three-month interval and approved for individuals aged 4 years and older, TAK-003 demonstrated moderate efficacy (∼61%) against virologically confirmed dengue infection in clinical trials, prompting its global rollout^11–13^.

Although efficacy trial studies reported higher neutralization titers in DENV-exposed individuals and consistently weaker responses to DENV-4, those studies concluded that TAK-003 was highly immunogenic across all groups, including DENV-naïve individuals^11–15^. However, these conclusions were primarily derived from double-blind, placebo-controlled efficacy trials settings, which may not reflect response magnitude, breadth, or underlying factors driving unbalanced immune responses. Importantly, the impact of immune imprinting and of the vaccine’s backbone on vaccine induced immune responses remain poorly defined. Given the rapid global implementation of Qdenga, a thorough understanding of its immunological profile is essential to prevent the recurrence of past vaccine hurdles. Here, we provide a detailed longitudinal characterization of humoral and cellular responses elicited by Qdenga in a real-world adult cohort from dengue-endemic regions, including older adults (aged 65–90+), a population excluded from initial clinical trials. We investigated whether the vaccine design skews responses to the DENV-2 backbone and assessed the extent to which immune imprinting (prior DENV exposure) shapes the magnitude and breadth of the vaccine responses.

## Results

### Immunological features of Qdenga vaccine

One hundred and ten participants from DENV-endemic regions in Brazil were enrolled in this longitudinal cohort study between August 2023 and June 2024 to evaluate immunological responses following vaccination. Consistent with the manufacturer’s recommendations, participants received two doses of Qdenga administered three months apart. Plasma and peripheral blood mononuclear cells (PBMCs) were collected at baseline (pre-vaccination), at the time of the second vaccination dose (90 days after the first dose), and at 7 and 60 days post-second dose **(Fig.1a).** Participants were stratified by sex, age, and prior DENV exposure. Pre-vaccination serostatus was used to infer prior DENV exposure, based on IgG DENV ELISA, with confirmation by neutralization assays using a titer cutoff of >1:20–1:40. Individuals seropositive for at least one serotype in both assays were classified as DENV-exposed, while those seronegative across all four serotypes were considered DENV-naive. This classification reflects the absence or presence of detectable DENV-specific humoral immunity at baseline. Cellular immune responses were characterized using flow cytometry and activation-induced marker (AIM) assays. Viral antibody serum levels and neutralization antibody activity against the four dengue vaccine strains (DENV-1, DENV-2, DENV-3, and DENV-4) were assessed using IgG ELISA and neutralization assays. Plaque titration and morphology for the four DENV strains are shown in the **Supplementary Fig. 1.** Basic demographic information, stratified by sex, age group and baseline serostatus, is provided in **Table S1.** Participants further completed questionnaires documenting prior flavivirus infections and provided proof of previous yellow fever vaccination, as detailed in **Table S1.** This information, along with baseline serological screening for yellow fever and Zika virus (ZIKV), was incorporated into the analysis to assess the potential confounding effects of pre-existing flavivirus immunity. The study cohort included 69 females (69.7%) and 30 males (30.3%). Although the study aimed for a balanced sex distribution, enrollment was based on voluntary participation, which led to a higher proportion of females. Participants were evenly distributed between younger adults (aged 18–64, 51.5%) and older adults (aged 65–90+, 48.5%). Half of the cohort (50%) presented serological evidence of previous dengue infection at baseline. Participants were monitored for local and systemic adverse events following each Qdenga dose through direct observation and structured follow-up questionnaires. Immediate post-vaccination reactions were recorded during a 30-minute clinical observation period, and delayed symptoms were tracked for up to three months. As summarized in **Table S2,** the most frequent events were pain at the injection site (11.3% after the first dose, 3.1% after the second) and transient systemic symptoms such as fever, malaise, or headache (<5% each). Most events resolved within 48 hours without medical intervention. One elderly participant experienced an acute coronary syndrome after the second dose; no causal association with vaccination could be established.

### Qdenga vaccination drives broad B cell activation and memory formation

The cellular mechanisms involved in dengue vaccine responses, including B cell activation and memory formation, remain poorly defined, as prior research has predominantly centered on serological outcomes^13,14,16,17^. To assess Qdenga cellular-induced immunity, we first longitudinally profiled total B and T cell populations as well as DENV-specific T cell responses following vaccination. Using PBMCs, we characterized total B cell dynamics over time, stratifying participants into two groups based on their baseline DENV serostatus (DENV-exposed and DENV-naïve). Overall, after Qdenga immunization, total B cell frequencies declined **(Supplementary Fig. 2a,b)**, suggesting a redistribution or differentiation of B cell subpopulations, as expected after vaccination. Early differentiated B cells (CD27−CD180+), essential for initiating immune responses, declined over time, with a more pronounced decrease in DENV-exposed individuals, likely reflecting a rapid recall from pre-existing memory pools **(Supplementary Fig. 2c,d)**. In contrast, multiple populations of activated B cells expanded following Qdenga vaccination **(Fig. 1b and Supplementary Fig. 2e-h)**. CD27+CD180+ B cells, associated with antigen-experienced and functionally responsive B cell subsets, increased progressively after vaccine doses in both groups **(Fig. 1b)**. Inflammatory B cells expressing HLA-DR and CXCR3, indicative of activated effector B cells capable of antigen presentation and tissue homing, also increased over time in both DENV-exposed and DENV-naive individuals **(Supplementary Fig. 2e,f)**. This increase suggests engagement of T cell-B cell interactions and memory formation. Additionally, we observed an increase in HLA-DR+CD86+ B cells, particularly in DENV-exposed individuals, with statistical significance reached post second vaccination dose **(Supplementary Fig. 2g,h)**. These mature, antigen-experienced cells subset also contribute to antibody production and memory formation. Total memory B cells (CD27+CD38−) and antibody-secreting cells (ASCs, CD27+CD38+) expanded gradually in both groups post-vaccination, with memory B cell increases more pronounced in seropositive individuals **(Fig. 1c,d)**. This shift was accompanied by a corresponding decline in naive B cell counterparts **(Supplementary Fig. 2i,j)**. To visualize global shifts in total B cell trajectory over time, we performed a combined analysis of activation and memory B cell subsets across all collection time points. This unsupervised analysis revealed a progressive transition in B cell phenotypes from baseline (T0) through successive post-vaccination time points (T1, T2, and T3) in peripheral blood, consistent with coordinated activation, differentiation, and memory formation following vaccine doses **(Fig. 1e)**. These findings demonstrate robust vaccine-induced total B-cell activation and phenotypic remodeling of the memory compartment, with broadly similar dynamics between groups but stronger responses among individuals with prior DENV exposure.

**Figure 1.**
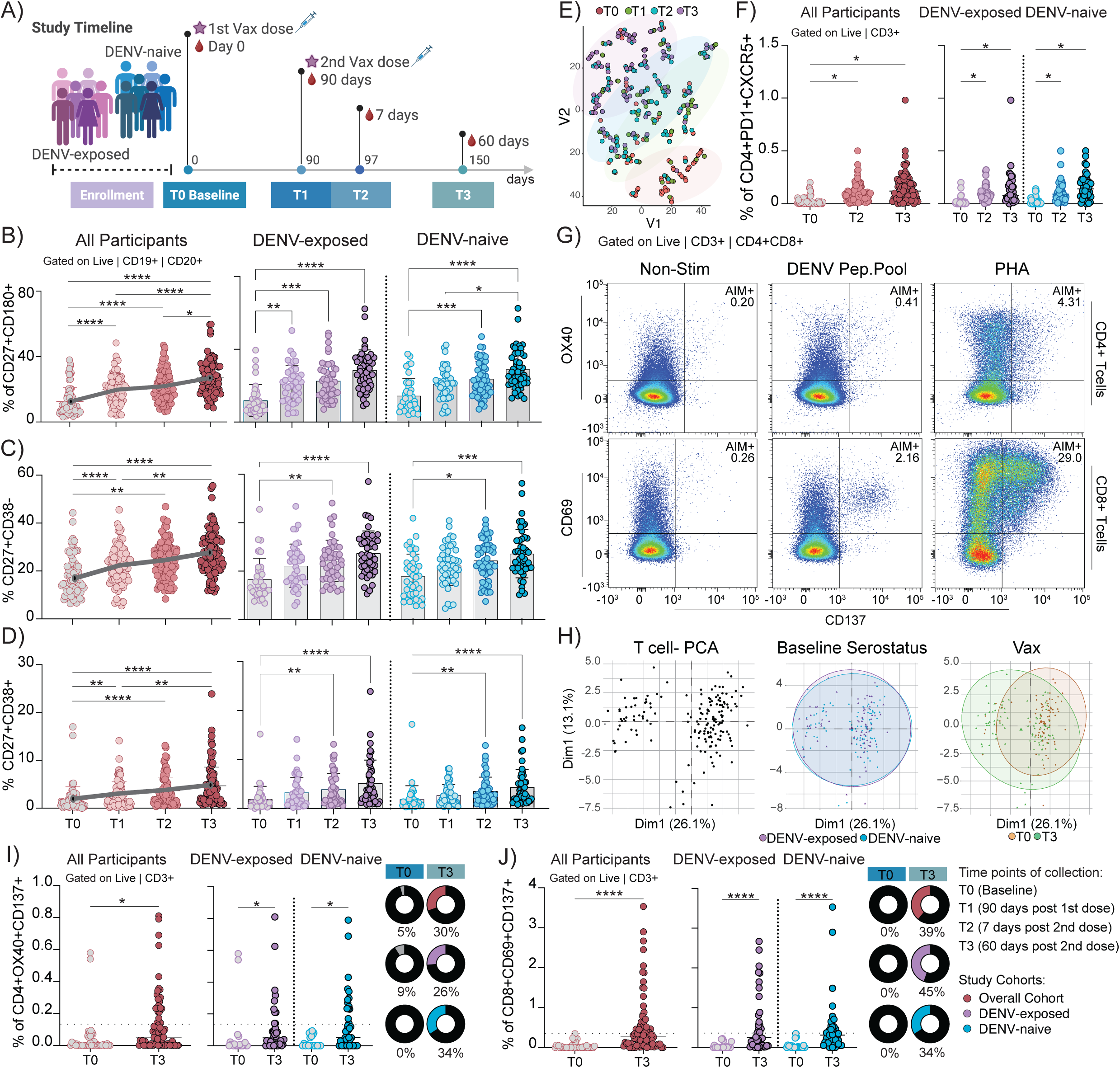
Characterization of cellular immune responses induced by Qdenga vaccination. **a,** Study timeline, participants received two doses of Qdenga at a 3-month interval. Plasma and PBMCs were collected at baseline (T0, prior to first dose), 90 days post-first dose/before the second dose (T1), 7 days post-second dose (T2), and 60 days post-second dose (T3). Participants were stratified by baseline serostatus: DENV-naïve (blue, n = 50) and DENV-exposed (purple, n = 49). Created with BioRender. **b-d,** Frequencies of B cell subsets over time in the full cohort (red, left) and stratified by baseline serostatus (right). B cell subsets were gated on singlets, live, CD19⁺CD20⁺ cells. **b,** CD27⁺CD180⁺ activated B cells; **c,** CD27⁺CD38⁻ memory B cells (MBCs); **d,** CD27⁺CD38⁺ antibody-secreting cells (ASCs). Significance was determined using the Kruskal–Wallis test followed by Dunn’s multiple comparisons test. Horizontal bars show mean values. **e,** t-distributed stochastic neighbor embedding (t-SNE) visualization of activation and memory B cell subsets across time points T0 (n = 74), T1 (n = 86), T2 (n = 94), and T3 (n = 80). Dots are color coded based on timepoints. Outlines were added to delineate cluster boundaries. **f,** Frequencies of circulating T follicular helper (Tfh) cells (CD3⁺CD4⁺PD-1⁺CXCR5⁺) in participants at T0 (n = 77), T2 (n = 92), and T3 (n = 80) in the full cohort (red, left) and stratified by baseline serostatus: DENV-naïve (blue, T0: n = 40; T2: n = 48; T3: n. = 40) and DENV-exposed (purple, T0: n = 37; T2: n = 44; T3: n. = 40). Significance was determined using the Kruskal–Wallis test followed by Dunn’s multiple comparisons test. **g-j,** DENV-reactive T cell responses following stimulation with virus-specific peptide pools. **g,** Representative gating strategy and dot plots for AIM⁺ CD4⁺ (OX40⁺CD137⁺) and AIM⁺ CD8⁺ (CD69⁺CD137⁺) T cells. **h,** Principal component analysis (PCA) of T cell memory subsets showing global distribution (left), baseline-driven distribution (middle), and vaccine-driven clustering (right) at T3. Data are projected onto PC1 (21.6% variance) and PC2 (13.1% variance). Ellipses represent 95% confidence intervals. Baseline, T0 (N = 77). T3 (N =92). **i–j,** Frequencies of AIM⁺ CD4⁺ (i) and AIM⁺ CD8⁺ (j) T cells in participants at T0 (n = 44), and T3 (n = 79) in the full cohort (red, left) and stratified by baseline serostatus: DENV-naïve (blue, T0: n = 20; T3: n. = 41) and DENV-exposed (purple, T0: n = 22; T3: n. = 39). Statistical significance was assessed using a two-tailed Mann–Whitney test. To control for background activation, frequencies from DENV-stimulated wells were normalized by subtracting paired unstimulated control values. Circular plots indicate the percentage of individuals above the positivity threshold (3 SD above baseline (dashed line). Each dot represents an individual; horizontal bars show median. Stim, non-stimulated cells; PHA, phytohemagglutinin was used as positive control. Flow-cytometry gating was used to identify and quantify distinct B and T cell populations within peripheral blood mononuclear cells. Each dot in the plots represents an individual cell; the clusters enclosed by gates (boxes) correspond to the subpopulation of interest, and the percentage displayed indicates its frequency within the parent population. ****P < 0.0001, ***P < 0.001, **P < 0.01, and *P < 0.05.

### DENV-specific T Cell responses induced by Qdenga across serostatus groups

Although T cell responses have recently been suggested as key correlates of protection against DENV infection^18,19^, virus-specific T cell responses following dengue vaccination remain largely uncharacterized. To investigate the T cell-mediated component of Qdenga-induced immunity and assess global T cell dynamics following vaccination, we analyzed DENV-specific T cell responses in our cohort using flow cytometry. First, we conducted longitudinal profiling of total CD4+ and CD8+ T cell subsets at baseline (prior to vaccination), 7 and 60 days post-second vaccination dose. As expected, vaccination was associated with a significant decrease in naïve T cell frequencies in both CD4+ and CD8+ compartments, accompanied by an increase in central memory T cells (TCM) **(Supplementary Fig. 3a,b).** In the overall CD8⁺ compartment, effector memory T cells (TEM) showed a trend toward increased frequency, although changes were not statistically significant **(Supplementary Fig. 3b).** We also observed a decrease in CD4+ and CD8+ T cells expressing PD-1 and TIM-3, two markers associated with T cell exhaustion, suggesting a potential shift toward a less exhausted^20,21^, more functionally responsive phenotype post-vaccination **(Supplementary Fig. 3c,d).** Of note, T follicular helper (Tfh) cells significantly expanded post-vaccination, potentially indicating increased T cell support for B cell activation **(Fig. 1f).** Overall, T cell phenotypic dynamics post-vaccination was similar between DENV-naive and DENV-exposed individuals. To assess virus-specific responses, T cells were evaluated for their ability to recognize DENV antigens through CD4 and CD8 specific peptide pools. Responses were assessed using activation-induced marker (AIM) assays, as previously described^19,22–24^. To detect virus-specific CD4+ and CD8+ T cell populations, PBMCs from vaccinated individuals were stimulated for 24h with the respective peptide pools at baseline and 60 days post second vaccination dose. Qdenga vaccination led to an increase in DENV-specific CD4+ and CD8+ T cells, as evidenced by the upregulation of activation markers, including OX40, CD69, and CD137 **(Fig. 1g,i,j)**. In addition, principal component analysis (PCA) of the T cell dataset revealed that virus-specific T cell profiles were not driven by baseline serostatus, age or sex. Instead, vaccination was the primary driver of cellular response clustering **(Fig. 1h and Supplementary Fig. 3e,f).** While increases in DENV-specific T cell responses were observed in both CD4⁺ and CD8⁺ compartments, the response was more pronounced in CD8⁺ T cells **(Fig. 1j).** Overall, 30% of participants exhibited detectable CD4⁺ T cell responses, while 39% showed CD8⁺ T cell reactivity post-vaccination **(Fig. 1i,j)**. These responses were comparable between DENV-exposed and DENV-naive individuals, indicating that prior DENV exposure did not significantly influence the magnitude of T cell responses post vaccination. The arithmetic means (95% CI) of DENV-specific CD4⁺ and CD8⁺ T cells in DENV-naïve individuals were 0.138% (95% CI: 0.081–0.196) and 0.465% (95% CI: 0.244–0.686), respectively; while in DENV-exposed participants they were 0.113% (95% CI: 0.055–0.170) and 0.591% (95% CI: 0.342–0.840) at day 60 post-vaccination. To further characterize the memory phenotype of these antigen-specific T cells, we assessed memory subset distribution within the AIM+ population. We observed an increase in central memory CD8⁺ T cells and effector memory CD4⁺ and CD8⁺ T cells among DENV-specific populations post-vaccination **(Supplementary Fig. 3g,h).** Thus, our data indicate that Qdenga vaccination effectively elicits virus-specific T cell responses independent of age, sex and baseline serostatus.

### Unsupervised analysis reveals serostatus- and age-driven neutralization profiles

To provide a complete view of Qdenga-induced immunity, we next assessed humoral responses. We first characterized neutralizing antibody responses using computational analyses, to provide an unbiased overview of vaccine-elicited humoral immunity, without imposing predefined groupings. Unsupervised hierarchical clustering was performed based solely on neutralization titers against all four DENV serotypes across all collected time points, spanning from baseline to 60 days post-second vaccine dose. This analysis identified five distinct participant clusters, each exhibiting unique neutralization profiles **(Fig. 2a, b).** Clusters 1 and 2 comprised participants with high neutralization titers. Participants on Cluster 4 displayed intermediate neutralization capacity, whereas participants on clusters 3 and 5 exhibited overall low neutralizing antibody levels post vaccination **(Fig. 2b).** To further delineate these clusters, we next assessed demographic and clinical cohort characteristics, including sex, baseline DENV serostatus and age. Although the cohort had an overall predominance of female participants **(Table S1)**, sex distribution was similar across all clusters **(Fig. 2c).** However, marked differences were observed regarding serostatus: clusters 1 and 2 primarily comprised individuals previously exposed to DENV (96% and 71% seropositive, respectively), whereas clusters 3, 4, and 5 predominantly included DENV seronegative individuals (87%, 69%, and 95%, respectively) **(Fig. 2d).** To complement this clustering analysis, a separate heatmap stratified by baseline serostatus further indicates the higher and broader neutralization responses in DENV-exposed individuals compared with DENV-naïve participants **(Supplementary Fig. 4a).** Age stratification revealed that clusters 1, 2, and 5 contained comparable proportions of younger (18–64 years) and older adults (65–90+ years). Notably, cluster 3 was predominantly composed of younger adults (73%), while cluster 4 included mostly older adults (85%) **(Fig. 2e).** These observations suggest that previous DENV exposure is strongly associated with higher neutralizing antibody levels post-vaccination, independent of age, as seen in clusters 1 and 2. Conversely, dengue-naïve participants consistently exhibited lower neutralization responses. Within this group, age influenced response magnitude (cluster 4), with older adults exhibiting high neutralization titers **(Fig. 2b,e).** Using random forest analysis, we further confirmed that, in contrast to T cell responses, baseline DENV serostatus and participants age were primary determinants of neutralization capacity post-Qdenga vaccination in our cohort **(Fig. 2f).** Other demographic factors, such as sex and prior yellow fever vaccination, were not significantly associated with differences in neutralization profiles (**Fig. 2c,e).**

**Figure 2.**
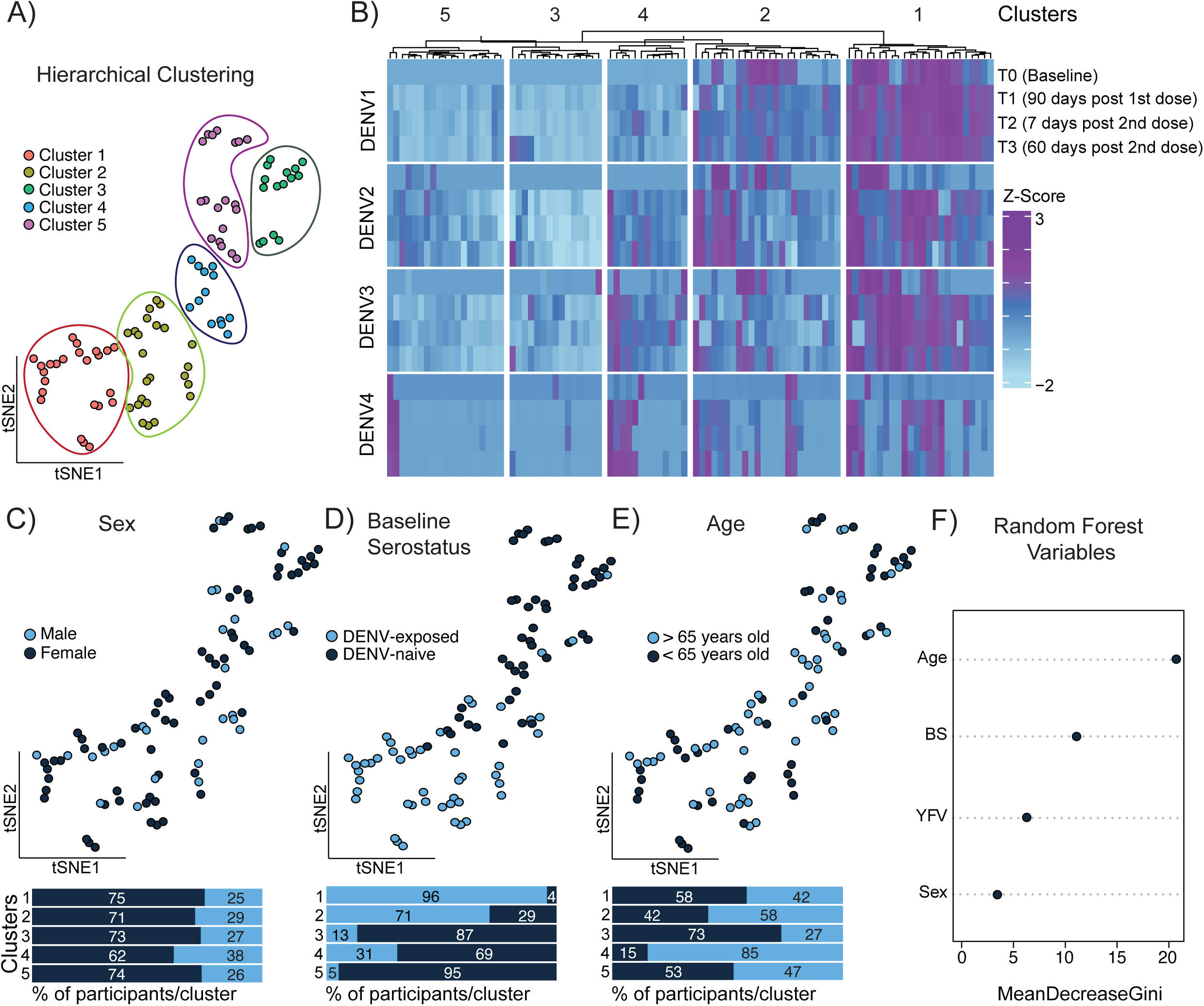
Neutralization profile-based clustering of Qdenga vaccinees. Plasma samples were collected from baseline to 5 months post-vaccination, and neutralization assays were performed against authentic DENV-1–4 using focus reduction neutralization tests (FRNT). **a,** t-SNE visualization of neutralization profiles against all four DENV serotypes across all time points (N = 95). Hierarchical clustering was performed using Euclidean distances. The number of clusters was defined by a prominent dissimilarity gap in the dendrogram. Outlines were added to delineate cluster boundaries. **b,** Heatmap of neutralization profiles across serotypes and timepoints for individual patients. Columns represent patient-specific FRNT50 responses, z-score normalized per serotype and timepoint. Darker blue tones indicate higher relative neutralization capacity._Hierarchical clustering (Euclidean distance) is shown in the upper dendrogram. **c–e,** Color-coded t-SNE distribution of participant characteristics across the five identified clusters: **c,** sex, **d,** baseline DENV serostatus, and **e,** age group. Bars indicate the percentage of participants for each feature within each cluster, with exact values indicated inside the bars. **f,** Random forest based analysis of demographic and immunological variables importance (age, baseline serostatus (BS), prior yellow fever vaccination (YFV), and sex) in hierarchical clustering. Mean Decrease Gini is a measure of variable importance. It quantifies how much a variable contributes to reducing node impurity (Gini impurity) across the trees in the forest. Higher values indicate greater importance in classifying the outcome.

### Neutralization kinetics indicates serostatus-dependent and serotype-skewed responses

To build upon the computational analysis and further define the temporal dynamics of vaccine-induced humoral immunity, we next evaluated antibody binding and neutralizing responses longitudinally. Given the impact of the DENV-baseline serostatus on humoral responses, participants were initially stratified into DENV-exposed and DENV-naïve groups **(Supplementary Fig.4b and Fig. 3a)**. Antibody binding was assessed by ELISA using the whole-virion (DENV-1–DENV-4). Overall, Qdenga vaccination elicited detectable anti-DENV IgG responses, with higher response rates to DENV-1 and DENV-2 (53-93%), followed by DENV-3 (44-89%), and lower binding to DENV-4 (8-44%), after the full vaccination regimen. However, antibody levels were significantly lower in DENV-naïve participants than those observed in previously exposed individuals **(Supplementary Fig.4b)**. We next assessed neutralizing capacity over time using a focus reduction neutralization test (FRNT), against the same DENV strains used in the Qdenga vaccine formulation^25^. Consistent with antibody binding analysis, neutralization titers among seronegative individuals were significantly lower across all DENV serotypes at all post-vaccination time points compared to seropositive participants **(Fig. 3a).** The only exception was DENV-2 at 60 days post-second vaccination dose, where no statistically significant difference was observed between groups **(Fig. 3a).** Of note, in DENV-naïve participants, neutralizing antibody responses were low across all serotypes, and no significant increase in DENV-4 neutralizing antibody titers was detected at any time point post-vaccination compared to baseline **(Supplementary Fig.5a,b).** Geometric mean titers (GMTs) for each DENV serotype, stratified by baseline serostatus and time point (T0–T3), are summarized in **Table S 3.** In contrast, seropositive individuals mounted higher neutralizing titers overall, with a skewed distribution favoring DENV-1, followed by DENV-3, DENV-2, and the lowest responses to DENV-4 **(Supplementary Fig.5a,b).** This pattern mirrors the baseline serotype reactivity and reflects regional virus circulation patterns^26^, as most previously DENV-exposed participants in our cohort had higher pre-vaccination titers against DENV-1 **(Supplementary Fig.5a).** The strong DENV-3 responses likely reflected cross-reactivity with DENV-1, given their close antigenic and structural similarity^27^, whereas DENV-2 responses corresponded to the vaccine backbone. Notably, peak neutralization capacity occurred after the first vaccine dose in both groups, with no further enhancement following the second dose **(Fig. 2b and Supplementary Fig.5a).** No significant differences were observed when stratifying the neutralization responses over time by sex groups across any of the four DENV serotypes **(Supplementary Fig. 5c).**

**Figure 3.**
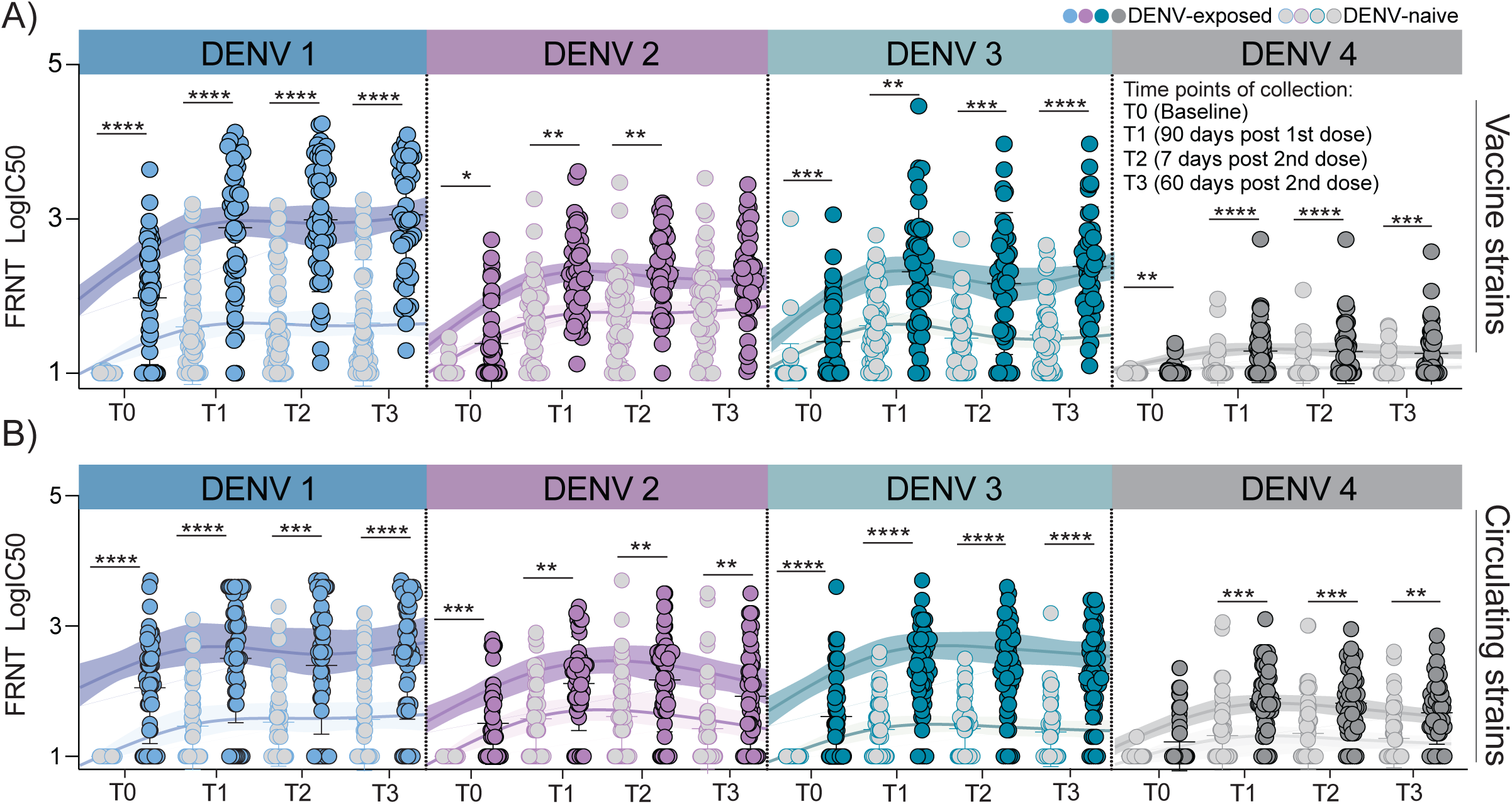
Temporal dynamics of neutralizing antibody responses against vaccinal and circulating DENV strains following Qdenga vaccination. Plasma samples were collected at baseline (T0, prior to first dose), 90 days post-first dose/before the second dose (T1), 7 days post-second dose (T2), and 60 days post-second dose (T3). Participants received two doses of Qdenga at a 3-month interval and were stratified by baseline serostatus: DENV-naïve (n = 50) and DENV-exposed (n = 49). **a,** Neutralization titers against the four vaccine DENV serotypes were quantified across all time points using a focus reduction neutralization test (FRNT) and stratified by baseline serostatus. **b,** Neutralization titers against circulating DENV strains stratified by baseline serostatus. Data are presented for all time points (T0–T3) with DENV-exposed individuals shown in full color and DENV-naïve individuals in lighter shades. Regression lines depict neutralization kinetics over time and are displayed in matching full or lighter shades to indicate group trends; shaded areas represent 95% confidence intervals. Horizontal bars indicate median values. Significance was determined using the Kruskal–Wallis test followed by Dunn’s multiple comparisons test. ****P < 0.0001, ***P < 0.001, **P < 0.01, and *P < 0.05.

Furthermore, to explore whether the observed neutralization patterns were specific to the vaccine strains or extended to naturally circulating viruses, we conducted parallel FRNT assays using contemporary DENV isolates representative of those currently circulating in Brazil. These assays revealed similar serostatus- and serotype-dependent trends, confirming that the immunodominance hierarchy observed for the vaccinal strains was preserved when tested against endemic viruses **(Fig. 3b).** These findings indicate that vaccine-induced antibodies recognize conserved epitopes shared across strains. In addition to neutralizing antibody activity, antibody Fc-mediated effector functions, including antibody-dependent complement deposition (ADCD), have recently been associated with protective immunity against dengue^28^. To evaluate whether complement enhances vaccine-induced antibody function, we performed neutralization assays in the presence of exogenous complement at 60 days post second vaccination dose. However, complement did not significantly increase neutralizing capacity in Qdenga-vaccinated individuals (**Supplementary Fig.5d).** Finally, since prior exposure to other flaviviruses such as ZIKV and yellow fever virus could act as a potential confounder, we measured baseline ZIKV and yellow fever virus neutralization capacity in all participants. Approximately 11% of participants had detectable anti-ZIKV neutralization titers at baseline, however, no significant differences were observed between the DENV-naïve and DENV-exposed groups (**Supplementary Fig. 5e)**. Similarly, neutralizing responses against the yellow fever vaccinal strain did not differ significantly across these groups (**Supplementary Fig. 5f).** These data suggest that Zika and yellow fever virus exposure did not impact neutralization responses elicited by Qdenga. Overall, these findings demonstrate that Qdenga-induced antibody responses are primarily shaped by prior DENV exposure, with limited enhancement after the second vaccination dose. Responses in DENV-naïve individuals were low and neutralization against DENV-4 was poor across both DENV serostatus groups.

### Antibody response is shaped by pre-vaccination immune imprinting in DENV-seropositives and vaccine backbone in seronegatives

Given the high variability in baseline neutralization titers, particularly among seropositive individuals, we normalized post-vaccination responses to each participant’s baseline to more accurately quantify vaccine-induced responses regardless of pre-existing immunity. Consistent with our initial clustering analysis **(Fig. 2)**, participants in Clusters 1 and 2 (composed predominantly of participants seropositive for DENV at baseline) exhibited the highest fold increases in neutralizing antibody titers following Qdenga vaccination **(Supplementary Fig. 6a).** Cluster 4 showed intermediate responses. Interestingly, although Cluster 5 (largely composed of DENV-naïve individuals) exhibited very low post-vaccination titers, after normalization, it demonstrated modest fold increases, particularly against DENV-2, the vaccine backbone **(Supplementary Fig. 6a).** This data indicates that individuals previously infected with DENV not only exhibit higher absolute antibody titers but also achieve greater increases in neutralization titers post-vaccination compared with seronegative individuals **(Supplementary Fig. 6a,b).** Specifically, DENV-exposed individuals exhibited significantly greater fold increases in neutralizing titers than DENV-naïve participants for DENV-1 (12.3× vs. 2.7×) and DENV-3 (9.0× vs. 2.8×), consistent with strong recall responses shaped by prior immunity. Of note, for DENV-2, both groups showed a boost in titers, and the difference was less pronounced (8.9× in DENV-exposed vs. 7.4× in DENV-naïve), reflecting the impact of the DENV-2 vaccine backbone. Minimal increases were observed for DENV-4 in both groups (1.22× in DENV-exposed vs. 1.0× in DENV-naïve), highlighting the consistently poor immunogenicity against this serotype **(Supplementary Fig. 6b).** Together, these results suggest that pre-existing immunity substantially amplifies vaccine responsiveness, while responses in naïve individuals are directed to the vaccine backbone and remain limited in magnitude and breadth.

Next, to assess population-level vaccine responsiveness based on neutralization capacity, we quantified the proportion of vaccine responders, defined as participants who exhibited a ≥0.5 log (3-fold) increases in neutralizing antibody levels relative to their own baseline. Overall, 76% of Qdenga-vaccinated participants mounted a neutralizing response to DENV-2, 63% to DENV-1, 61% to DENV-3, and only 12% to DENV-4 **(Fig. 4a).** These proportions remained stable across longitudinal time points, emphasizing the limited immunological impact of the second vaccination dose on neutralization capacity **(Fig. 4b).** Dengue vaccines are currently designed to induce protection against all four DENV serotypes; therefore, antibody breadth is a key immunological metric of their tetravalent immunogenicity. In this study, breadth was defined as the number of DENV serotypes (out of four) for which an individual mounted a neutralizing response, determined by a ≥0.5 log (3-fold) increase from to their own baseline titers. Participants were subsequently classified into five categories: non-responders (0 serotypes), narrow (1 serotype), limited (2 serotypes), broad (3 serotypes), and tetravalent (4 serotypes). Striking differences were observed between individuals with and without prior DENV exposure. Among DENV-exposed participants, 63% (46% broad, 17% tetravalent) mounted responses to at least three serotypes, with only 12% classified as non-responders **(Fig. 4c and Supplementary Fig. 7a).** In contrast, DENV-naïve individuals exhibited a narrower response profile: 27% responded to three serotypes, only 8% achieved tetravalent responses, and 18% failed to respond to any serotype. Additionally, nearly one-third (29%) of DENV-naïve participants mounted responses to only a single serotype **(Fig. 4c).** Among these narrow responders, the vast majority responded exclusively to DENV-2, which was the serotype present in the vaccine backbone **(Supplementary Fig. 7b).** Notably, even within the subset of broad responders in the DENV-naïve group, post-vaccination serotype dominance remained skewed toward DENV-2 **(Supplementary Fig. 7c).** These data demonstrate that prior DENV exposure not only enhances the magnitude **(Fig. 3b)** but also the breadth of vaccine-induced responses **(Fig. 4c).** Despite this, tetravalent responses remained limited across the cohort, with particularly low rates among DENV-naïve individuals. These findings highlight the persistent challenge in dengue vaccine development in achieving a uniformly robust tetravalent response, particularly in seronegative populations.

**Figure 4.**
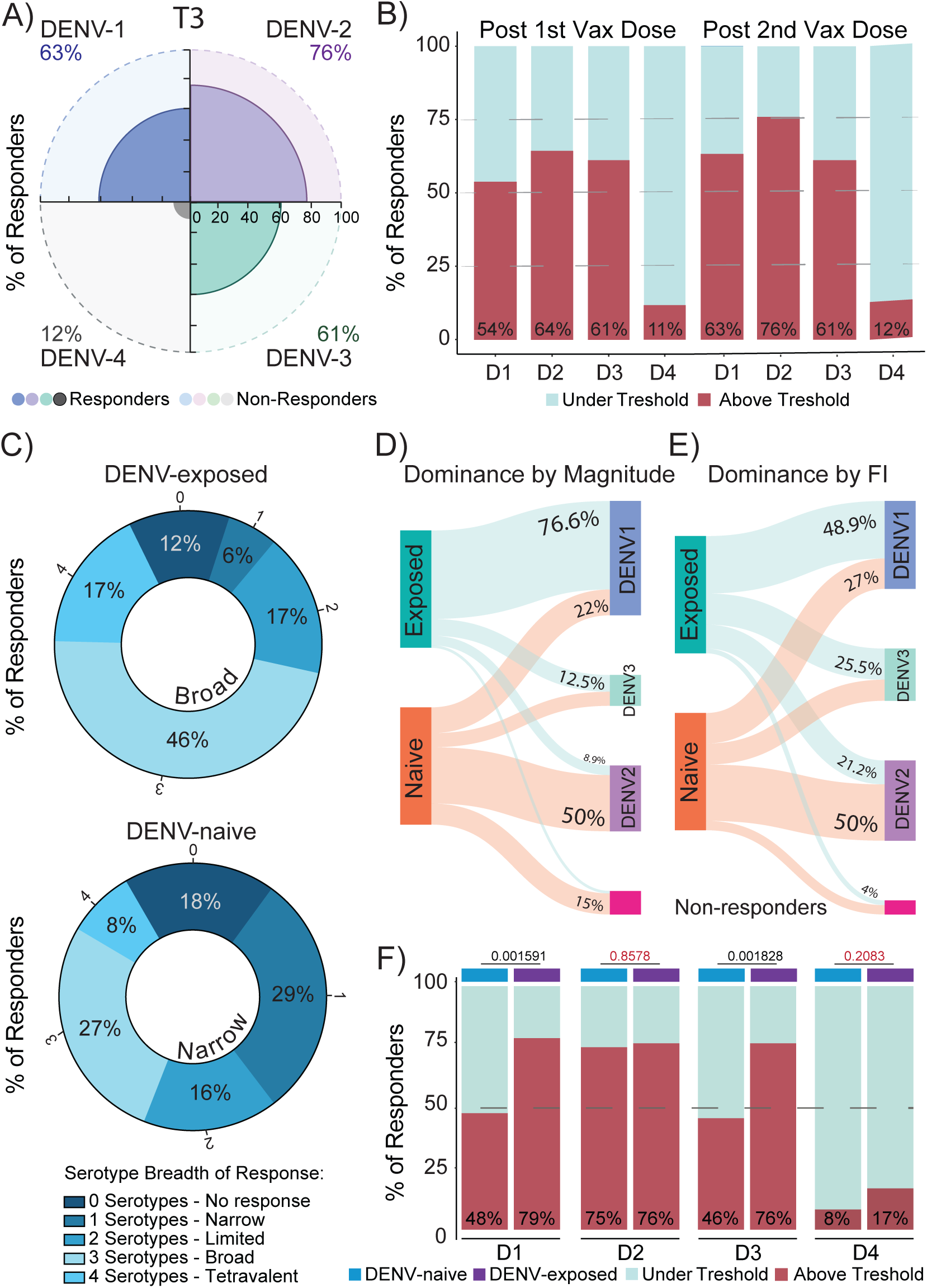
Responsiveness, breadth, and serotype dominance of humoral responses following Qdenga vaccination. **a,** Pie chart indicating the percentage of vaccine responders for each serotype at 60 days post-second dose (T3). Responders were defined as individuals who achieved a ≥0.5 log (3-fold) increase in neutralization titers relative to their baseline (T0). Data are shown for DENV-1 (blue), DENV-2 (purple), DENV-3 (green), and DENV-4 (gray). Solid bars represent responders; hatched bars indicate non-responders. **b,** Stacked bar plots representing the percentage of vaccine responders above (red) or below (light green) the 3-fold threshold after the first (T1) and second (T3) vaccination dose, for each of the four dengue virus serotypes (DENV-1 to DENV-4, denoted as D1–D4). **c,** Pie chart distribution of immunization breadth after full vaccination, stratified by baseline serostatus: DENV-exposed (top) and DENV-naïve (bottom). Individuals were categorized based on the number of DENV serotypes eliciting a neutralizing response: 0 (no response), 1 (narrow), 2 (limited), 3 (broad), or 4 (tetravalent). **d–e,** Serotype dominance after Qdenga vaccination (T3). **d,** Dominance by magnitude, defined as the serotype with the highest post-vaccination neutralization titer. **e,** Dominance by fold-increase (FI), defined as the serotype showing the greatest rise in neutralization titer relative to baseline. Sankey diagrams depict the shift in dominance patterns for DENV-exposed and DENV-naïve individuals; line widths indicate the percentage of participants per serotype. **f,** Stacked bar plots representing the percentage of participants achieving serotype-specific immunization threshold after the second vaccine dose (T3), stratified by baseline serostatus (DENV-exposed, DENV-naïve). Responders were defined using the same ≥0.5 log (3-fold) increase criterion described in panel a,b. Pairwise comparisons were performed using a nonparametric t-test to assess the impact of prior DENV exposure on vaccine-induced immunity for each serotype. Panels **d** and **e** were generated using SankeyMATIC (https://sankeymatic.com/) to visualize serotype dominance patterns. Participants were stratified by baseline serostatus: DENV-naïve (blue, n = 50) and DENV-exposed (purple, n = 49). Significance was determined using the Kruskal–Wallis test followed by Dunn’s multiple comparisons test. Exact p values are reported where applicable.

To better understand how baseline imprinting and vaccine composition shape the specificity of vaccine-induced responses, we next examined whether the dominant serotype response post-vaccination could be predicted by pre-existing serotype-specific antibody profiles. This analysis aimed to disentangle the contributions of immune imprinting and the potential influence of Qdenga’s DENV-2 backbone. We assessed serotype dominance in two complementary ways: (1) by magnitude, identifying the serotype with the highest neutralization titer post-vaccination; and (2) by fold increase, identifying the serotype with the greatest rise in neutralization titer relative to each participant’s own baseline. At baseline, 74.4% of DENV-exposed individuals exhibited the highest titers against DENV-1, with smaller subsets showing dominance for DENV-3 (15.4%) and DENV-2 (10.3%) **(Supplementary Fig. 7d).** This dominance pattern was largely retained post-vaccination, with 76.6% of DENV-exposed participants still exhibiting the strongest neutralizing responses against DENV-1, followed by DENV-3 (12.5%) and DENV-2 (8.4%) **(Fig. 4d).** DENV-naive participants had a distinct post-vaccination profile, with dominant responses primarily skewed toward DENV-2 (50%), followed by DENV-1 (22%) and DENV-3 (13%), respectively **(Fig. 4d).** When dominance was assessed by fold increase from baseline, accounting for pre-existing immunity, similar trends emerged. Among DENV-exposed individuals, responses were most frequently directed toward DENV-1 (48.9%), followed by DENV-3 (25.5%) and DENV-2 (21.2%). In DENV-naïve individuals, fold-dominant responses remained centered on DENV-2 (50%), followed by DENV-1 (27%) and DENV-3 (18%) **(Fig. 4e).** Furthermore, when stratifying vaccine responders by baseline serostatus, we further confirmed the role of immune imprinting and vaccine composition in shaping population-level responsiveness. As expected, the proportion of responders that reached the threshold 3-fold (≥0.5 log10) for DENV-1 and DENV-3, was significantly higher among participants who were seropositive at baseline. However, no significance in responder rates was observed for DENV-2 between seropositive and seronegative individuals. Of note, for DENV-4, no difference in vaccine responsiveness was detected between groups; however, this was due to uniformly low responder rates across both seropositive and seronegative individuals **(Fig. 4f).** These data indicate that serotype-specific immune imprinting plays a dominant role in shaping vaccine responses among previously DENV-exposed individuals, with Qdenga largely reinforcing pre-existing antibody hierarchies. In contrast, in DENV-naïve individuals, responses were predominantly directed toward DENV-2, reflecting the influence of the vaccine’s DENV-2 backbone. These results highlight that both prior immunity and vaccine design are the main drivers contributing to serotype dominance post-vaccination.

### Qdenga elicits immune responses in the elderly

Given that individuals aged 65 and older were not included in TAK-003’s efficacy trials, the immunogenicity and effectiveness of the vaccine in this population remain unclear. Additionally, in our cohort, age and baseline DENV serostatus emerged as primary determinants of post-vaccination neutralizing antibody responses **(Fig. 2f).** To further investigate the influence of age on vaccine-induced immunity, we stratified participants by baseline DENV serostatus and age group and analyzed neutralization magnitude, fold change, and antibody breadth following Qdenga administration. Although our study was not powered to evaluate safety outcomes, no differences in reactogenicity were observed between younger and older participants, supporting the vaccine’s safety profile across age groups **(Table S 2).** Of note, one elderly participant experienced an acute coronary syndrome two days after the second dose; however no direct causal association with vaccination could be established. While neutralization responses among seropositive individuals were comparable between age groups, marked age-related differences emerged within the seronegative cohort **(Fig. 2b,e).** Longitudinal analysis of neutralization kinetics post vaccination further supported these findings, showing that the pronounced differences between seropositive and seronegative groups were primarily driven by younger individuals **(Supplementary Fig. 8a).** This pattern was consistent across fold increases from baseline, vaccine responsiveness and antibody breadth **(Supplementary Fig. 8b-d).** Remarkably, older seronegative individuals (≥65 years) demonstrated intermediate neutralization responses, higher than those of younger seronegatives but lower than seropositive participants **(Supplementary Fig. 8a,b).** Of note, the serological profiles, by magnitude and fold increase of older seronegative individuals more closely related to those of seropositive participants than their younger seronegative counterparts. However, unlike seropositives, responses in aged individuals were predominantly directed against DENV-2, aligning with the vaccine’s backbone **(Supplementary Fig. 8e).** To investigate potential mechanisms underlying the enhanced vaccine responsiveness observed in seronegative older adults, we first evaluated prior exposure to other circulating flaviviruses in Brazil, such as ZIKV and yellow fever virus. Neutralization assays performed at baseline revealed no significant differences in anti-ZIKV or anti-YFV neutralizing antibody titers between older and younger individuals **(Supplementary Fig. 8f,g)**. We next assessed whether delayed clearance of vaccine-derived viremia in older seronegative participants could explain their heightened antibody responses, however, viremia was undetectable in both older and younger participants at 7 days post-second vaccination dose (data not shown). This analysis suggests no distinct patterns or prolonged viral persistence based on age, suggesting that neither prior flavivirus exposure nor differential control of vaccine viremia accounts for the age-related differences in neutralizing antibody responses. Together, these findings demonstrate that Qdenga is well-tolerated in older adults and induces both neutralizing antibody and T cell responses in DENV-exposed and DENV-naïve individuals. However, although baseline DENV-naïve older adults mounted higher responses than their younger counterparts, their responses remained narrower and predominantly DENV-2–focused compared to those of seropositive participants.

Finally, to examine whether cellular responses might compensate for limited antibody responses, we next correlated AIM⁺ T-cell data with neutralization breadth profiles. Participants were categorized according to their antibody-response breadth (no response, narrow (1serotype), limited (2 serotypes), broad (3 serotypes), or tetravalent (4 serotypes) and analyzed for the presence of DENV-reactive CD4⁺ and CD8⁺ T-cell activation (**Supplementary Fig. 7e**). The frequency of participants exhibiting activation of dengue-specific CD4⁺, CD8⁺, or combined CD4⁺/CD8⁺ T-cell responses was comparable across all antibody-response categories. Notably, individuals with absent or narrow antibody responses did not exhibit higher frequencies of virus-specific T-cell activation relative to those with broader or tetravalent neutralization, suggesting no inverse relationship between humoral and cellular responses. We next characterized the “best responders” within our cohort, defined as individuals who (1) achieved neutralizing responses (≥0.5 log increase) to at least three DENV serotypes and (2) ranked among the top 30% for DENV-1, DENV-2, and DENV-3 neutralizing antibody levels at 60 days post-second dose. Over 80% of these individuals exhibited detectable virus-specific CD4⁺ and CD8⁺ T cell responses, suggesting coordinated humoral and cellular immunity. No significant differences were observed in this group based on sex, age, or markers of prior flavivirus exposure, including baseline ZIKV neutralization titers and history of yellow fever vaccination. Prior DENV exposure was predominant, with over 85% of best responders being seropositive at baseline, highlighting pre-existing immunity as the key factor in achieving broad and high-magnitude responses to Qdenga **(Supplementary Fig. 7f).** Together, these findings indicate that Qdenga vaccination does not elicit balanced neutralizing antibody responses, but rather serostatus-dependent and backbone-skewed immunity. Prior DENV exposure enhances both the magnitude and breadth of vaccine-induced response, shaped by baseline serotype-specific imprinting. In contrast, seronegative individuals have lower neutralization responses overall, with limited breadth and bias toward the vaccine backbone. Overall, these results emphasize the critical role of DENV immune history in shaping the magnitude, balance, and breadth of neutralizing responses to Qdenga vaccination.

## Discussion

In this study, we provide a comprehensive immunological assessment of the Qdenga vaccine in a real-world adult cohort from a dengue-endemic region of Brazil. By integrating longitudinal neutralization kinetics, antibody breadth, and paired B and T cell profiling, we addressed key gaps that have persisted since the vaccine global rollout. We demonstrated that the DENV-2 backbone of Qdenga drove type-2–biased responses in naïve individuals and failed to overcome pre-existing serotype imprinting in DENV-exposed participants. Overall, Qdenga does not elicit balanced neutralizing antibody responses across the four DENV serotypes, but rather serostatus-dependent and serotype-skewed immunity. Among participants with prior DENV exposure, serotype-dominant responses were predominantly shaped by baseline immunity, highlighting immune imprinting as the key determinant of response magnitude and breadth. In contrast, among baseline DENV-naïve individuals, we observed markedly lower responses skewed toward DENV-2, the vaccine backbone. These findings help clarify why achieving balanced tetravalent immunity remains challenging for Qdenga and identify both pre-existing immunity and the DENV2 backbone as the main drivers contributing to the incomplete and uneven serotype coverage observed in earlier trials. Of note, neutralizing antibody titers plateaued after the first vaccination, with no substantial increase following the second dose. We further demonstrated that Qdenga induced B and T cell responses, revealing broad cellular activation across serostatus groups and expansion of memory and effector populations. Collectively, these results fill several gaps left by previous studies and have important implications for vaccine design and deployment.

Our data confirm and expand upon some observations from Takeda’s clinical trials and immunogenicity studies, such as higher neutralizing antibody responses in DENV-exposed individuals and consistently weaker responses to DENV-4. However, our findings diverge from Takeda’s overall conclusion that TAK-003 is broadly immunogenic across all groups, including DENV-naïve participants^14,15,29^. This divergence stems mainly from differences in how immunogenicity was assessed. Previous studies relied on binary seropositivity thresholds (e.g., a 1:10 dilution cutoff) and geometric mean titers (GMTs), metrics commonly used in vaccine efficacy trials. For example, Sirivichayakul et al. reported 100% “positivity” to DENV-1–3 in adults 4 months post-vaccination, regardless of baseline serostatus^14^. In contrast, our analysis focused on response magnitude and breadth and was based on full neutralization curves and incorporated both group-level and intra-individual evaluations, allowing direct assessment of vaccine-induced increases and serotype dominance. Using this approach, we found that only 8% of DENV-naïve individuals achieved tetravalent responses, while nearly one-third responded to a single serotype (predominantly DENV-2). Additionally, at group-level analysis, DENV-naïve participants exhibited no significant increase in DENV-4 neutralization titers at any time point relative to baseline, raising important concerns about the level of protection achievable in naïve populations. Although clinical outcomes such as ADE or breakthrough infections were not assessed in this study, our findings highlight the need for close monitoring of vaccine effectiveness during outbreaks, particularly in DENV-naïve regions.

We also showed that DENV-2 backbone–driven dominance was largely restricted to seronegative individuals, while vaccine responses in seropositive participants were primarily shaped by pre-existing immunity. In our cohort, 74.4% of DENV-exposed individuals exhibited baseline dominance for DENV-1^26^, a pattern largely maintained post-vaccination (76.6%). This indicates that Qdenga predominantly reinforces existing serotype hierarchies rather than reshaping them. We speculate that immune imprinting in participants with baseline DENV-1 dominance may act synergistically with the DENV-2 backbone to expand breadth, as reflected by the broader responses observed in seropositive individuals. However, due to insufficient numbers of individuals with baseline dominance for DENV-2-4, we were unable to assess whether these findings were generalizable across serotypes. Future studies designed to stratify participants by baseline serotype dominance will be critical to fully elucidate how imprinting modulates the specificity and breadth of vaccine-induced response.

Our analysis of total B cell dynamics revealed progressive increases in activated and memory B- cell subsets. The observed B-cell remodeling following Qdenga vaccination resembles the kinetics described after natural secondary DENV infection, characterized by decreased of circulating early-differentiated B cells and expansion of antigen-experienced subsets^30^. The increase in HLA-DR⁺CXCR3⁺ B cells is consistent with effective vaccine-induced activation rather than pathogenic inflammation, supporting a protective immune profile^31,32^. Together, these findings suggest that Qdenga triggers coordinated and functional B-cell activation. However, our analysis was limited to total B cell phenotypes, and virus-specific memory B cells were not assessed. Future studies using antigen-specific probes will be necessary to characterize the quality and longevity of vaccine-induced memory B cells, particularly in seronegative populations with lower neutralizing antibody responses. Additionally, although complement-mediated neutralization has been associated to protection in dengue infection^28^, the addition of complement did not further enhance neutralizing activity in our assays, suggesting that Qdenga-elicited antibodies primarily mediate protection through direct virion neutralization rather than Fc-dependent mechanisms. In contrast to antibody responses, DENV-specific T cell responses expanded across all groups regardless of age, sex, or baseline serostatus. This is consistent with prior findings in pediatric and adolescent cohorts showing that TAK-003 induces DENV-specific CD4⁺ and CD8⁺ T cell responses independent of serostatus^33^. While our assays capture T cell reactivity rather than direct measures of protection, the balanced activation of CD4⁺ and the more pronounced responses among CD8⁺ AIM⁺ T cells observed here align with studies in both human and non-human studies, where DENV-specific T cells have been linked to viral clearance and protection from severe disease^24,32,34^. Notably, only a subset of participants (∼35%) displayed detectable DENV-reactive T cells activation, indicative of modest cellular engagement overall. Additional studies will be required to assess the durability, effector capacity, and potential protective contribution of these responses.

Older adults (≥65 years) were excluded from TAK-003’s safety, immunogenicity, and efficacy trials, leaving a critical gap in understanding vaccine performance in this population. We anticipated reduced immunogenicity, as observed with other viral vaccines^35–40^, but instead found that older adults mounted detectable humoral and cellular responses. In this cohort, seropositive older adults performed comparably to their younger counterparts, while seronegative older adults showed intermediate humoral responses, higher in magnitude than younger seronegatives but still biased toward DENV-2. No significant differences were observed in B or T cell responses by age. This unexpected finding may potentially reflect lifelong exposure to multiple flaviviruses in endemic regions such as Brazil, potentially priming the immune system despite undetectable antibody levels at baseline. It is plausible that prior flavivirus exposures occurred long ago or repeatedly, with waning antibody titers below detection thresholds, particularly relevant given evidence of faster antibody decay in the elderly for other pathogens^35,41,42^. However, in our cohort, prior flavivirus exposure, assessed by neutralization antibody titers against yellow fever and Zika virus, did not differ between younger and older adults at baseline. Future studies incorporating virus-specific memory B cell assays could provide a more sensitive measure of prior flavivirus encounters at the cellular level. Additionally, we also investigated whether elderly participants might exhibit prolonged vaccine-derived viremia, previously reported with yellow fever vaccination, which could potentially contribute to stronger antibody responses^43^. However, our data showed no differences in viremia clearance between younger and older participants at 7 days post-second dose. We acknowledge that this time point may miss earlier dynamics, and analysis immediately following the first vaccine dose would be more informative. Overall, our findings suggest that Qdenga is immunogenic in older adults, including those without prior DENV exposure, although the breadth of these responses remains limited in this group. In our cohort, Qdenga was generally well tolerated across age groups and serostatus categories. Most local and systemic reactions were mild and self-limiting, consistent with previous TAK-003 safety reports^12^. Importantly, one previously DENV-exposed participant over 70 years old experienced an acute coronary syndrome two days after the second vaccination dose; while no direct causal relationship with vaccination could be established, this event highlights the importance for continued post-vaccination monitoring, particularly in older adults or individuals with possible underlying cardiovascular conditions. It is important to note that this cohort was established primarily to evaluate vaccine-induced immunogenicity rather than safety outcomes; while adverse events were systematically monitored and reported, the study was not powered to assess clinical safety or detect rare adverse events.

Taken together, our study provides immunological insights into the performance of the Qdenga vaccine. The combination of vaccine backbone–driven skewing in naïve individuals and immune imprinting in previously exposed individuals limits both the magnitude and breadth of neutralizing antibody responses, which helps to explain why balanced tetravalent immunity is rarely achieved. These findings raise important considerations for vaccine deployment: the influence of immune imprinting, where prior DENV exposure shapes the hierarchy of vaccine-induced serotype responses, suggests that immunological history must be considered when evaluating vaccine performance. Moreover, the consistently poor response to DENV-4 across groups raises concerns about the completeness of tetravalent coverage, a critical goal for dengue vaccines. Given that the second Qdenga dose showed limited impact on enhancing either antibody titers or cellular responses, these findings support the potential need for booster doses using reformulated vaccines designed specifically to improve DENV-4 immunogenicity. Notably, the Butantan TV003/TV005 vaccine has demonstrated stronger responses against DENV-4 in clinical trials^17^, and future studies could explore whether heterologous boosting with such candidates could rescue the poor DENV-4 responses observed with Qdenga. These findings not only improve our understanding of Qdenga’s immunological profile but also offer actionable insights to optimize dengue vaccination strategies across diverse populations, particularly in endemic regions facing ongoing transmission and shifting serotype prevalence.

## Data Availability

All data produced in the present study are available upon reasonable request to the corresponding author.

## Methods

### Ethic and inclusion statement

This study was approved by the National Research Bioethics Committee of Brazil CAAE 74062523.4.0000.5149. Written informed consent was obtained from all enrolled participants prior to study procedures. Clinical and demographic data were collected using REDCap 9.3.6. Participants were enrolled between August 2023 and June 2024. Basic demographic information, stratified by sex, age group and baseline serostatus, is provided in **Table S1.** No serious adverse events were reported following Qdenga vaccination **(Table S2).** Although adverse events were monitored and reported, the cohort was primarily established for immunogenicity analyses rather than for safety surveillance and was not powered to detect rare outcomes.

### Study Participants

A total of 110 participants from Minas Gerais, Brazil, were enrolled in this longitudinal cohort study to evaluate immunological responses following Qdenga vaccination. Eleven individuals did not return for study visits after vaccination and were therefore excluded, resulting in a final study cohort of 99 participants. Participants received two doses of Qdenga (lot: 543916) administered three months apart, following manufacturer recommendations. Plasma and peripheral blood mononuclear cells (PBMCs) were collected at four time points: at baseline (pre-vaccination), at the time of the second vaccination dose (90 days after the first dose), and at 7 and 60 days post-second dose **(Fig.1a).** Participants were stratified by sex, age, and prior dengue virus (DENV) exposure. Pre-vaccination serostatus was determined using a combination of commercial DENV IgG ELISA (Abbott, Ref. 01PE30), in-house whole-virion ELISA, and focus reduction neutralization tests (FRNT). Participants who tested positive in at least two of these assays were classified as DENV-exposed, while those negative for all four serotypes were considered DENV-naïve. Demographic information, prior flavivirus infection history, and yellow fever vaccination status were collected using questionnaires and confirmed, when possible, with vaccination records. Additional baseline serological screening for yellow fever virus (YFV) and Zika virus (ZIKV) was performed to control for potential confounders. Although this cohort was established primarily to characterize vaccine-induced humoral and cellular immunity, participants were also monitored for clinical safety and reactogenicity. Local and systemic symptoms were recorded by clinical staff during a 30-minute observation period after each dose and through standardized follow-up questionnaires for three months post-vaccination. However, the study was not powered to evaluate safety outcomes or detect rare adverse events. Detailed clinical and demographic data, including sex and age distribution (younger adults: 18–64 years; older adults: 65–90+ years), are provided in **Table S 1 and Table S 2**. Overall, the study cohort included 69 females (69.7%) and 30 males (30.3%), with equal representation of younger and older adults (51,5% vs. 48.5%). Half of the cohort (50%) had evidence of prior DENV exposure at baseline. Due to limited PBMC availability, sample allocation was prioritized hierarchically across assays, first for antigen-specific AIM T-cell assays, followed by full B-cell phenotyping and, when sufficient cells remained, partial T-cell phenotyping. All ELISA, FRNT, and flow cytometry analyses were performed in a blinded manner by independent laboratory teams.

### Cell Lines and viruses

Vero cells (ATCC® CCL-81) were cultured in Dulbecco’s Modified Eagle Medium (DMEM; Gibco) supplemented with 10% fetal bovine serum (FBS) and 1% sodium pyruvate and maintained at 37 °C with 5% CO₂. Dengue virus serotypes DENV-1 (strain 16007), DENV-2 (strain 16681), DENV-3 (strain 16562), and DENV-4 (strain 1036) were kindly provided by Dr. Claire Huang (CDC) and expanded in Vero cells. Viral stocks were harvested at 6–7 days post-infection, clarified by centrifugation (450 × g for 5 min), filtered through 0.70 µm filters, and titrated by FRNT. Brazilian isolates of circulating DENV strains were also tested in neutralization assays: **DENV-1:** genotype V, accession no. PV682577, **DENV-2:** genotype IV, accession no. PV682585, **DENV-3:** genotype V, accession no. PV685005, **DENV-4:** genotype II, accession no. PV683387. Zika virus (ZIKV-BRA; accession no. KX280026) and yellow fever virus (YFV-17D; accession no. X03700) were similarly propagated in Vero cells. ZIKV viral stocks were harvested at 48 h post-infection, and YFV at 3 days post-infection. Both were clarified by centrifugation, filtered through 0.70 µm filters, and titrated by FRNT.

### Isolation of plasma and PBMCs

Whole blood was collected in heparinized CPT vacutainer tubes (BDAM362780, BD) and processed on the day of collection. Plasma was separated by centrifugation at 600 × g for 20 min at room temperature and stored at −80 °C in cryogenic vials (Sigma Aldrich, V7884-450EA). PBMCs were isolated according to the manufacturer’s instructions, washed twice with PBS, and counted after brief ACK lysis (2 min). Cell viability was determined by trypan blue exclusion using an automated cell counter (Thermo Fisher, AMQAX1000). PBMCs were cryopreserved at −80 °C for later analysis.

### Flow cytometry

PBMCs were thawed and stained using the following antibodies: BV605 anti-CD3 (UCHT1, 1:300, BioLegend), BV785 anti-CD4 (SK3, 1:300, BioLegend), BV421 anti-CCR7 (G043H7, 1:50, BioLegend), AF700 anti-CD45RA (HI100, 1:100, BioLegend), APC anti-CD69 (FN50, 1:100, BioLegend), BV711 anti-CD137 (4B4-1, 1:50, BioLegend), APC/Fire750 anti-CD8 (SK1, 1:200, BioLegend), PE anti-PD1 (EH12.2H7, 1:100, BioLegend), PE-CF594 anti-CD25 (567489, 1:200, BD Biosciences), FITC anti-CXCR5 (J252D4, 1:100, BioLegend), PE-Cy7 anti-TIM3 (F38-2E2, 1:100, BioLegend), BV785 anti-CD19 (SJ25C1, 1:300, BioLegend), BV421 anti-CD138 (MI15, 1:300, BioLegend), AlexaFluor700 anti-CD20 (2H7, 1:200, BioLegend), AlexaFluor647 anti-CD27 (M-T271, 1:350, BioLegend), PE/Dazzle594 anti-IgD (IA6-2, 1:400, BioLegend), and BV711 anti-CD38 (HIT2, 1:200, BioLegend). Cells were incubated with Live/Dead Fixable Aqua (Thermo Fisher) for 20 min at 4 °C, blocked with Human TruStain FcX (BioLegend), and stained for 30 min at room temperature. Secondary markers were applied for an additional 30 min at 4 °C. Samples were fixed with 4% paraformaldehyde (PFA) for 30 min, washed, and acquired on an Attune NXT flow cytometer (Thermo Fisher). Data were analyzed using FlowJo software v10.6 (TreeStar). The gating strategy used in this study is fully described in **Supplementary Figure 9.**

### T cell stimulation

DENV1-4 CD4 MegaPool (MP) and DENV1-4 CD8 MP are synthetic peptide pools developed based on experimentally validated DENV epitopes. These DENV-specific MPs were designed to combine a large number of peptides derived from all four dengue virus serotypes into single pools through sequential lyophilization approaches. The DENV1-4 CD4 and CD8 MPs used in this study consist of 180 and 268 peptides and include 15-mer and 9/10-mer peptides, respectively, as previously described in the literature^22^. Thawed PBMCs (5–8 × 10⁵ cells/well) were stimulated with DENV-specific CD4 and CD8 peptide megapools (2 µg/mL per peptide) for 24 h at 37 °C, 5% CO₂. On day 0, PBMCs were thawed, counted, and plated in a total of 5–8 × 105 cells per well in 200 ul of RPMI 1640 medium (Gibco) supplemented with 1% sodium pyruvate (NEAA), 100 U/ml penicillin–streptomycin (Biochrom) and 20% FBS at 37 °C and 5% CO2. On day 1, cells were washed, and the stimulation was performed with peptide megapools (MP) identified as: DENV CD4 and DENV CD8 as previously described^18^. Negative controls included unstimulated cells, and phytohemagglutinin (PHA) served as a positive control. After stimulation, cells were washed in PBS-2 mM EDTA and processed for flow cytometry. To control for background activation, frequencies from DENV-stimulated wells were normalized by subtracting paired unstimulated values on a per-individual basis. Vaccine responders were defined as individuals whose post-vaccination (T3) responses exceeded two standard deviations above their baseline (T0) frequency. AIM assays were restricted to baseline (T0) and 60 days after the second dose (T3) due to limited PBMC availability, capturing pre-vaccination and post-booster responses.

### Focus reduction neutralization test (FRNT)

Neutralization assays were performed as previously described^44^. Heat-inactivated plasma (56 °C for 30 min) was 3-fold serially diluted (1:20–1:43,740 for DENV and YFV; 1:20–1:5120 for ZIKV) and incubated with virus for 1 h at 37 °C. For DENV-1–4 and YFV-17D, the mixture was added to confluent Vero cells in 96-well plates for 1 h, followed by overlay with DMEM containing 2% FBS and 1% carboxymethylcellulose. Plates were incubated for 48 h (DENV-3 and DENV-4) or 60 h (DENV-1, DENV-2, and YFV), fixed with 10% formaldehyde, permeabilized (Invitrogen, #00-8333-56), and blocked using the same buffer with 5% non-fat dry milk for 10 min. Cells were stained with monoclonal antibodies 4G2 and 2H2 (1:700), followed by HRP-conjugated anti-mouse IgG (H+L) secondary antibody (SeraCare, #5220-0341; 1:1500) and developed with TrueBlue peroxidase substrate (SeraCare, #5510-0030). Plates were air-dried and read using an ImmunoSpot reader. Parallel assays were performed using the DENV strains included in the Qdenga vaccine formulation and contemporary circulating isolates collected in Brazil, allowing direct comparison of vaccine-strain and field-strain neutralization profiles.

FRNT assays were also performed in the presence of 7% baby rabbit complement (BioRad, C12CA.1) to evaluate complement-mediated neutralization. Rabbit complement was added 30 minutes prior to virus–plasma incubation and maintained during the 1-hour incubation at 37 °C. All subsequent steps, including infection, overlay, fixation, staining, and quantification, followed the standard FRNT protocol. We tested a range of complement concentrations (2%–10%) to determine the optimal amount that did not block viral entry or interfere with the neutralization assay. Both direct and complement-mediated neutralization assays were always performed with the respective viral controls, using established viral concentrations to generate 60–120 foci per well.

For ZIKV neutralization, the FRNT was performed in 24-well plates with Vero cells at ∼90% confluence. Virus inoculum (50–100 FFU/well) was incubated with 3-fold serially diluted plasma in MEM with 2% FBS and antibiotics for 1 h at 37 °C. After incubation, 180 µL of the virus–plasma mixture was added to the cells and incubated for another hour. Wells were overlaid with 1.6% carboxymethylcellulose in 2× MEM with 2% FBS and antibiotics and incubated for 3 days. Cells were fixed with methanol/acetone (1:1), blocked with PBS + 3% FBS, and stained overnight with anti-ZIKV primary antibody (UTMB, WRCEVA). HRP-conjugated goat anti-mouse IgG secondary antibody (SeraCare, #5220-0341) and TrueBlue substrate were used for signal detection. Plates were air-dried and read using an ImmunoSpot reader.

### Whole-Virion ELISA

To measure dengue-specific binding antibodies, high-binding ELISA plates (Corning, Ref. 3690) were coated overnight at 4 °C with DENV virions (1 × 10^5^ PFU/plate) in 0.1 M carbonate buffer (Thermo Scientific, Cat. J62610.AP). The following day, plates were washed once with 1× TBS-T (Cell Signaling, Ref. 9997S) and then blocked with blocking buffer (1× TBS-T supplemented with 5% non-fat dry milk) for 2h at room temperature.

Plasma samples were diluted 1:600 in blocking buffer and added in duplicate to the wells (30 μL per well). Plates were incubated overnight at 4 °C with the plasma samples. The next day, plates were washed three times with 1× TBS-T. A horseradish peroxidase (HRP)-conjugated anti-human IgG secondary antibody (SouthernBiotech, Cat. 2081-05) was diluted 1:4000 in blocking buffer and added to each well (30 μL per well) for 2 hours at room temperature. Plates were developed with TMB substrate (Thermo Scientific, Ref. 34028) and read at 450 nm.

### Statistical analysis

All statistical analyses were performed using GraphPad Prism 10, JMP 15, Python, R 4.3.1, and SankeyMatic (https://sankeymatic.com). Multiple group comparisons were assessed using the Kruskal–Wallis test with false discovery rate (FDR) correction for multiple comparisons. Pairwise comparisons were performed using the Mann–Whitney test. Flow cytometry analysis was performed using FlowJo 10.10.0. Participants were classified as responders if they exhibited a ≥3-fold increase in FRNT₅₀ titers at T1, T2, or T3 relative to their baseline (T0). Serotype dominance was defined as the DENV serotype with the highest FRNT₅₀ titer (log10) at each visit.

All statistical analyses were performed as specified in each figure legend and Results subsection. Both group-level and individual-level analyses were used throughout the manuscript, depending on the scientific question addressed. Group-level analyses were conducted to evaluate overall cohort trends and to compare post-vaccination timepoints against baseline (T0). These comparisons were performed using non-parametric tests (Kruskal–Wallis with Dunn’s multiple-comparisons correction) or, when longitudinal alignment of repeated measures was required, using mixed-effects models (REML, matched by subject and allowing for missing values). This approach captures population-level changes in immune parameters. Individual-level analyses were employed to account for inter-individual variability driven by pre-existing immunity and to evaluate vaccine-induced responses relative to each participant’s own baseline. For these analyses, post-vaccination values were normalized to baseline titers, yielding fold-change ratios (e.g. T3/T0). Participants were classified as responders if their post-vaccination neutralization titers exhibited at least a ≥3-fold increase compared with baseline, a cutoff that exceeds intra- and inter-assay variation and lies well above the 95% confidence range of assay noise. This combined framework, using both cohort-level statistics and participant-normalized fold-change analyses, allowed us to capture broad population trends while also dissecting the influence of pre-existing DENV immunity on individual vaccine responsiveness.

## Acknowledgements

We are very grateful to all study participants who kindly donated specimens for this study. We also would like to thank D. Mucida who provided insight and expertise that greatly assisted the data analysis and comments that significantly improved this manuscript. We extend our sincere gratitude to the sample processing, coordination and clinical support team, Lisa Esper, Josiane Frattari and Fatima Brant, for their invaluable role in establishing the study platform and their dedicated commitment to engaging with the study participant. We also thank Mônica Mayumi Akinaga, Bianca Gabrielly Silva Calbo, Natalia Franco Bueno Mistrão, MSc, and Milene Rocha Ribeiro, PhD, for their technical assistance. Additionally, we warmly thank Sophia Peng for her dedicated technical support, thoughtful discussions, and the enthusiasm she brought to our lab during her rotation. This study was supported by Stavros Niarchos Foundation Institute for Global Infectious Disease and by (NIAID/NIH) grant P01 AI106695.

## Author contributions

Conceived the study: MMT, CL.

Coordinated sample collection, processing, and shipping: FT, PRJA, MMT.

Performed virus sequencing: MIB, NDG.

Conducted cell culture and virus expansion: FA, LC.

Provided virus and assistance with virus-related protocols: JP, DRM, CH.

Performed Zika virus and circulating DENV neutralization assays: TMR, ABO, MLN.

Performed Yellow fever neutralization assays: FA.

Performed experiments and collected data: LC, VSM, JF.

Computational analysis: JF, AP.

Provided experimental support: JC, LL.

Assisted with peptide pool design and protocols: DW.

Analyzed data: LC, VSM, JF, CL.

Wrote the manuscript: LC, CL.

Supervised the project: CL.

All authors reviewed and approved the final manuscript.

## Materials and Data availability

All the background information of participants and data generated in this study are included in Source Data Figure1. The genome information and aligned consensus genomes for DENV used in this study are available on NCBI (GenBank Accession numbers: **DENV 1:** genotype II (AF180818), **DENV 2:** genotype Asian I (U87412), **DENV 3:** genotype V (KU725665) and **DENV 4:** genotype IIb (U18429) for DENV-vaccine strains. **DENV-1:** genotype V (PV682577), **DENV-2:** genotype IV (PV682585), **DENV-3:** genotype V (PV685005), **DENV-4:** genotype II (PV683387) for circulating DENV isolates. All data analysis used in this study is available at **Source data** file. Additional correspondence and requests for materials should be addressed to the corresponding author.

## Supplementary Figures

**Table S1. Cohort Demographics.** Clinical and demographic data for the participants enrolled in the Qdenga Study. Information is presented for the overall cohort and stratified by baseline DENV serostatus into DENV-naïve and DENV-exposed groups. For each category, total numbers are provided alongside the corresponding percentage of participants (shown in parentheses). Immunization breadth was defined based on the number of DENV serotypes eliciting a detectable neutralizing antibody response post-vaccination: 0 = no response; 1 = narrow response; 2 = limited response; 3 = broad response; 4 = tetravalent response.

**Table S2. Reactogenicity profile following Qdenga vaccination.** Summary of local and systemic symptoms observed or reported following the first and second doses of Qdenga. Immediate post-vaccination reactions were directly observed by clinical professionals during a 30-minute monitoring period, while subsequent events were self-reported by participants through structured questionnaires. Monitored symptoms included local reactions and systemic manifestations. Events were graded by intensity (mild, moderate, severe). Exact two-sided 95% confidence intervals were calculated using the Clopper–Pearson method for binomial proportions. The figure presents three panels: overall cohort (all participants), participants stratified by age group (<65 years, ≥65 years), and participants stratified by baseline DENV serostatus (DENV-naïve and DENV-exposed).

**Table S3. Geometric mean neutralizing antibody titers (GMTs) against DENV-1–4 vaccinal strains following Qdenga vaccination.** Neutralization was measured by focus reduction neutralization test (FRNT) using plasma collected at baseline (T0), at the time of the second vaccination dose (90 days after the first dose, T1), and at 7 days (T2) and 60 days (T3) post-second dose. Participants were stratified by baseline dengue serostatus (DENV-naïve and DENV-exposed). Data are shown as GMTs with 95% confidence intervals (CIs) for each DENV serotype.

**Supplementary Data Figure 1: Representative FRNT neutralization curves.** Representative plaque morphology for each DENV serotype used in the focus reduction neutralization assay (FRNT). Titration curves from a representative DENV-seropositive vaccinated individual at 60 days post-second Qdenga dose. Neutralization curves are shown for all four vaccine DENV serotypes (DENV-1–4). Participants were stratified by baseline serostatus: DENV-naïve (blue, n = 50) and DENV-exposed (purple, n = 49). Plasma was serially diluted in 8 threefold dilution steps, ranging from 1:20 to 1:43,720.

**Supplementary Data Figure 2. Characterization of B cell subpopulations dynamics following Qdenga vaccination.** Frequencies and gating strategies for major B cell subsets gated on live, single CD19⁺CD20⁺ cells. Samples were collected at four time points: T0 (baseline, prior to first dose), T1 (90 days post-first dose/before the second dose), T2 (7 days post-second dose), and T3 (60 days post-second dose). Participants were stratified by baseline serostatus: DENV-naïve (blue, n = 47) and DENV-exposed (purple, n = 45). Frequencies and representative plots of **(a,b)** total B cells, **(c,d)** early differentiated B cells (CD27⁻CD180⁺), **(e,f)** inflammatory B cells (CXCR3⁺HLA-DR⁺), **(g,h)** activated B cells (HLA-DR⁺CD86⁺), and **(I,j)** naïve B cells (CD27⁻CD38⁻) across all participants (red, left) and stratified by baseline serostatus: DENV-naïve (blue) and DENV-exposed (purple). Percentages for each gated population are indicated on the plots. Antibody-secreting cells, ASCs. Memory B cells, MBCs. Significance was determined using the Kruskal–Wallis test followed by Dunn’s multiple comparisons test. Flow-cytometry gating was used to identify and quantify distinct B cell populations within peripheral blood mononuclear cells. Each dot in the plots represents an individual cell; the clusters enclosed by gates (boxes) correspond to the subpopulation of interest, and the percentage displayed indicates its frequency within the parent population. ****P < 0.0001, ***P < 0.001, **P < 0.01, and *P < 0.05.

**Supplementary Data Figure 3. Characterization of T cell subpopulations induced by Qdenga vaccination. a– b**. Frequencies of CD4⁺ **(a)** and CD8⁺ **(b)** T cell subsets defined by CCR7 and CD45RA expression: naïve (CCR7⁺CD45RA⁺), central memory (TCM, CCR7⁺CD45RA⁻), effector memory (TEM, CCR7⁻CD45RA⁻), and terminally differentiated effector memory (TEMRA, CCR7⁻CD45RA⁺). Data are shown at baseline (T0, prior to first dose, n = 77), 7 days post-second dose (T2, n = 92), and 60 days post-second dose (T3, n = 80). **c–d**. Frequency of CD4⁺PD1⁺TIM3⁺ **(c)** and CD8⁺PD1⁺TIM3⁺ **(d)** T cells at T0 and T3, stratified by baseline serostatus: DENV-naïve (blue, T0: n = 40; T2: n = 48; T3: n. = 40) and DENV-exposed (purple, T0: n = 37; T2: n = 44; T3: n. = 40). Significance was determined using the Kruskal–Wallis test followed by Dunn’s multiple comparisons test. **e–f,** Principal component analysis (PCA) of total T cell subsets showing no clustering by **(e)** age or **(f)** sex at T3. **g–h.** DENV-reactive T cell responses following stimulation with PrM and E peptide pools. Frequencies and distribution of AIM⁺ T cell subpopulations at T0 and T3: **(g)** AIM⁺ CD4⁺ T cells (OX40⁺CD137⁺) and **(h)** AIM⁺ CD8⁺ T cells (CD69⁺CD137⁺) further stratified by CCR7 and CD45RA expression into naïve, TCM, TEM, and TEMRA subsets stratified by baseline serostatus: DENV-naïve (blue, T0: n = 20; T3: n. = 41) and DENV-exposed (purple, T0: n = 22; T3: n. = 39). Statistical significance was assessed using a two-tailed Mann–Whitney test. Data are stratified by baseline serostatus: DENV-exposed (purple, V0, n = 49 V2, n = 49 V3, n = 49) and DENV-naïve (blue, n = 50) groups. Flow-cytometry gating was used to identify and quantify distinct T cell populations within peripheral blood mononuclear cells. Each dot in the plots represents an individual cell; the clusters enclosed by gates (boxes) correspond to the subpopulation of interest, and the percentage displayed indicates its frequency within the parent population.****P < 0.0001, ***P < 0.001, **P < 0.01, and *P < 0.05.

**Supplementary Data Figure 4. Neutralization profile and antibody binding induced by Qdenga vaccination.** Plasma samples were collected at four time points: baseline (T0, prior to first dose), 90 days post-first dose/before the second dose (T1), 7 days post-second dose (T2), and 60 days post-second dose (T3). **a,** Heatmap of neutralization profiles (FRNT₅₀) against vaccine DENV strains across all four timepoints (T₀–T₃), separated by baseline DENV serostatus: DENV-naive (left) and DENV-exposed (right) participants. Columns represent patient-specific FRNT50 responses, z-score normalized per serotype and timepoint. **b,** Plasma IgG binding reactivity against DENV-1–4 was assessed by whole-virus ELISA (1:600 serum dilution, OD 450 nm) using vaccine strains at all time points. Each plate was coated with 1 × 10^5^ PFU of the corresponding virus.

**Supplementary Data Figure 5. Dynamics of neutralizing antibody responses induced by Qdenga vaccination a-c,** Neutralization titers against DENV-1–4 vaccine strains across all time points post-vaccination. Plasma samples were collected at four time points: baseline (T0, prior to first dose), 90 days post-first dose/before the second dose (T1), 7 days post-second dose (T2), and 60 days post-second dose (T3). **a,** Neutralization titers at all time points in DENV-naïve individuals (top panel) and DENV-exposed individuals (bottom panel). **b,** Comparison of neutralization titers at T3 (60 days post-second dose) across all four DENV serotypes, shown separately for DENV-naïve (top) and DENV-exposed (bottom) participants. **c,** Neutralization titers across all time points, stratified by sex: females (full color) and males (grey). Significance was determined using the Kruskal–Wallis test followed by Dunn’s multiple comparisons test. **d,** Neutralization titers against DENV-2 at T3 in the presence or absence of exogenous complement (n = 40 participants). **e,** Neutralization titers against Zika virus at baseline (T0) stratified by baseline DENV serostatus (n = 97 participants). **f,** Neutralization titers against yellow fever virus strain 17D at baseline (T0) in individuals with confirmed yellow fever vaccination, stratified by baseline DENV serostatus (n = 38 participants). Significance was determined using Mann–Whitney test. ZIKV, Zika virus. YFV, yellow fever virus. ****P < 0.0001, ***P < 0.001, and *P < 0.05.

**Supplementary Data Figure 6. Differential fold-change in neutralization titers across DENV serotypes. a,** Unsupervised heatmap displaying fold changes in neutralization titers (post-vaccination relative to baseline) across the four vaccine DENV serotypes (n=95). Columns represent individual responses; hierarchical clustering (Euclidean distance) is shown in the upper dendrogram. Neutralization titers at each post-vaccination timepoint were expressed as fold-change relative to each participant’s baseline (T0) FRNT value for the corresponding DENV serotype (post-TX / baseline-T0). **b,** Paired analysis of neutralization titers (FRNT LogIC50) at baseline (T0) and 60 days post-second dose (T3) for each DENV serotype. Results are stratified by baseline serostatus: DENV-naïve (left) and DENV-exposed (right). Each gray line represents an individual participant’s response; the colored line indicates the mean trajectory for each group. Fold-change values (T3/T0) are shown above each panel.

**Supplementary Data Figure 7. Breadth and serotype dominance of vaccinated cohort. a,** Pie chart depicting baseline serostatus distribution among individuals achieving the immunization threshold for ≥3 serotypes. DENV-exposed (blue) and DENV-naïve (white). **b,** Treemap representation of dominant serotype response among individuals achieving the immunization threshold for only one serotype, stratified by baseline serostatus. **c,** Treemap representation of dominant serotype response among individuals achieving the immunization threshold for only one serotype, stratified by baseline serostatus. DENV-exposed (left, dark) and DENV-naïve (right, light). Responders were defined as individuals achieving a ≥0.5 log (3-fold) increase in neutralization titers relative to their own baseline (T0). Only serotypes for which participants met the defined immunization threshold (≥3-fold increase from baseline) are represented **d,** Pie chart depicting baseline DENV serotype distribution among seropositive individuals (DENV-exposed). **e,** Alluvial plots showing the distribution of DENV-reactive CD4⁺ and CD8⁺ AIM⁺ T-cell responses across participants stratified by antibody-response breadth (no response, narrow (1 serotype), limited (2 serotypes), broad (3 serotypes), and tetravalent (4 serotypes). The width of each flow indicates the relative frequency of participants within each category, shown as percentages. T-cell responses were measured 60 days after the second vaccine dose and compared with corresponding antibody-response profiles (n = 79). Positive T-cell responses were defined as AIM⁺ frequencies above background-subtracted unstimulated controls and exceeding the predefined positivity threshold described in the Methods section. **f,** Stacked bar plots showing demographic characteristics (DENV baseline serostatus, sex, age group, hierarchical cluster assignment, yellow fever vaccination history, ZIKV baseline serostatus, breadth of response, and serotype dominance), of top responders. Best responders were defined as individuals achieving the immunization threshold for more than 3 serotypes and ranking among the top 30 FRNT50 titers for DENV-1, 2 and 3 (n = 14).

**Supplementary Data Figure 8. Impact of age on Qdenga immunogenicity.** Plasma samples were collected at baseline (T0, prior to first dose), 90 days post-first dose/before the second dose (T1), 7 days post-second dose (T2), and 60 days post-second dose (T3). Participants received two doses of Qdenga at a 3-month interval and were stratified by age group, adults (20–64 years) and elderly (≥65 years), and by DENV baseline serostatus. **a,** Neutralization antibody levels against the four vaccine DENV serotypes (DENV-1 to DENV-4) were quantified across all time points using a focus reduction neutralization test (FRNT). **b,** Mean fold increase in neutralization titers at T3 for DENV-1 to DENV-4, stratified by age and serostatus: DENV-naïve adults (black), DENV-exposed adults (gray), DENV-naïve elderly (dark gray), and DENV-exposed elderly (light gray). **c,** Stacked bar plots representing the percentage of vaccine responders for each serotype at T3. Responders were defined as individuals who achieved a ≥0.5 log (3-fold) increase in neutralization titers relative to T0. Data shown for DENV-1 (blue), DENV-2 (purple), DENV-3 (green), and DENV-4 (gray). Age groups are denoted in light brown (adults) and dark brown (elderly). **d,** Pie chart distribution of immunization breadth after full vaccination, stratified by age and baseline serostatus. Individuals were categorized based on the number of DENV serotypes eliciting a neutralizing response: 0 (no response), 1 (narrow), 2 (limited), 3 (broad), or 4 (tetravalent). **e,** Percentage of individuals exhibiting serotype dominance at T3, defined as the serotype with the highest neutralization titer, stratified by age and serostatus. **f,** Neutralization titers against Zika virus at T0, stratified by age and DENV serostatus (n = 97 participants). **g,** Neutralization titers against yellow fever virus strain 17D at T0 in individuals with confirmed yellow fever vaccination, stratified by age and DENV serostatus (n = 38 participants). ZIKV, Zika virus. YFV, yellow fever virus. n = 26 DENV-naïve adults, n = 24 DENV-naïve elderly, n = 25 DENV-exposed adults, and n = 24 DENV-exposed elderly. Regression lines depict neutralization kinetics over time; shaded areas represent 95% confidence intervals. Statistical analysis was performed using the non-parametric Kruskal–Wallis test followed by Dunn’s multiple comparisons test. ****P < 0.0001, ***P < 0.001, **P < 0.01, and *P < 0.05.

**Supplementary Data Fig. 9. Gating strategies for B and T cell subpopulations. a–d,** Representative flow-cytometry gating strategy used to define major T- and B-cell subsets from peripheral blood mononuclear cells (PBMCs). Lymphocytes were first identified based on **(a)** forward- and side-scatter parameters (FSC-A/SSC-A), **(b)** followed by exclusion of doublets using SSC-H vs SSC-W and **(c)** FSC-H vs FSC-W, and **(d)** selection of live cells using a viability dye. **e–i, B cell populations.** Total B cells were defined as CD19⁺CD20⁺ **(e).** Within this population, the following subpopulations were identified: (**f,** bottom right) early-differentiated B cells (CD19⁺CD20⁺CD27⁻CD180⁺); (**f,** top right) antigen-experienced B cells (CD19⁺CD20⁺CD27⁺CD180⁺); **(g)** activated B cells (CD19⁺CD20⁺HLA-DR⁺CD86⁺); (**h,** bottom left) naïve B cells (CD19⁺CD20⁺CD27⁻CD38⁻); (**h,** bottom right) memory B cells (MBC; CD19⁺CD20⁺CD27⁺CD38⁻); (**h,** top right) antibody-secreting cells (ASC; CD19⁺CD20⁺CD27⁺CD38⁺); and (i) inflammatory B cells (CD19⁺CD20⁺HLA-DR⁺CXCR3⁺). **j–q, T cell populations. (j)** T cells were first gated on live CD3⁺ single cells (viability dye-negative; CD3⁺Aqua⁻). CD4⁺ and CD8⁺ subsets were then defined **(k),** and within each, functional populations were delineated as follows: **l,** circulating T follicular helper cells (cTfh; CD4⁺PD-1⁺CXCR5⁺); **m,** exhausted T cells (TIM-3⁺PD-1⁺); **n,** naïve T cells (CCR7⁺CD45RA⁺), central memory (TCM; CCR7⁺CD45RA⁻), effector memory (TEM; CCR7⁻CD45RA⁻), and terminally differentiated effector memory cells (TEMRA; CCR7⁻CD45RA⁺). In each plot, each dot represents a single cell, and the gated populations of interest are outlined by rectangular regions. The percentage displayed within each gate represents the frequency of that subset relative to its parent population.

