## Supplementary Table S1 for "Serotype skewing and immune imprinting shape response to the tetravalent dengue virus Qdenga vaccine"

| | Overall (N = 99) | DENV-naive (N = 50) | DENV-exposed (N = 49) |
|---|---|---|---|
| **Age Category** | | | |
| 18 - 64 years | 51 (51.5) | 26 (52) | 25 (51) |
| 65 - 99 years | 48 (48.5) | 24 (48) | 24 (49) |
| **Sex** | | | |
| Female | 69 (69.7) | 35 (70) | 34 (69.4) |
| Male | 30 (30.3) | 15 (30) | 15 (30.6) |
| **ZIKV Serostatus at baseline** | | | |
| Negative | 86 (88.6) | 47 (94) | 39 (83) |
| Positive | 11 (11.4) | 3 (6) | 8 (17) |
| **Yellow fever vaccine (Decade of Vaccination)** | | | |
| 2020s | 1 (1.01) | 1 (2) | 0 |
| 2010s | 23 (23.3) | 10 (20) | 13 (26.5) |
| 2000s | 8 (8.06) | 6 (12) | 2 (4.08) |
| 1990s | 6 (6.03) | 3 (6) | 3 (6.12) |
| Unknown / No record | 61 (61.6) | 30 (60) | 31 (63.3) |
| **Breadth of Response** | | | |
| No response | 15 (15.8) | 9 (18.7) | 6 (12.8) |
| Narrow | 17 (17.9) | 14 (29.2) | 3 (6.4) |
| Limited | 16 (16.8) | 8 (16.7) | 8 (17.0) |
| Broad | 35 (36.8) | 13 (27.1) | 22 (46.8) |
| Tetravalent | 12 (12.6) | 4 (8.3) | 8 (17.0) |

Suppl. Table 1
