## Supplementary Table S2 for "Serotype skewing and immune imprinting shape response to the tetravalent dengue virus Qdenga vaccine"

| All Particpants | | |
|---|---|---|
| Event | After 1st dose<br>% (n, CI 95% Clopper-Pearson) | After 2st dose<br>% (n, CI 95% Clopper-Pearson) |
| **Local reactions** (at the application site) | | |
| Pain | 11.3% (11, 95% CI 5.8%–19.4%) | 3.1% (3, 95% CI 0.6%–8.8%) |
| Hematoma/Ecchymosis/Hypersensitivity | 1.0% (1, 95% CI 0.0%–5.6%) | 3.1% (3, 95% CI 0.6%–8.8%) |
| Erythema/Redness | – | 2.1% (2, 95% CI 0.3%–7.3%) |
| **Systemic** | | |
| Fever | 3.1% (3, 95% CI 0.6%–8.8%) | 2.1% (2, 95% CI 0.3%–7.3%) |
| Nausea | 3.1% (3, 95% CI 0.6%–8.8%) | - |
| Flu-like ilness | 2.1% (2, 95% CI 0.3%–7.3%) | 3.1% (3, 95% CI 0.6%–8.8%) |
| Diarrhea | 2.1% (2, 95% CI 0.3%–7.3%) | – |
| Malaise/Fatigue/Myalgia | 1.0% (1, 95% CI 0.0%–5.6%) | 3.1% (3, 95% CI 0.6%–8.8%) |
| Headache | – | 2.1% (2, 95% CI 0.3%–7.3%) |
| Retro-orbital pain | 1.0% (1, 95% CI 0.0%–5.6%) | 1.0% (1, 95% CI 0.0%–5.6%) |
| Chest pain | 1.0% (1, 95% CI 0.0%–5.6%) | – |
| Acute coronary syndrome | – | 1.0% (1, 95% CI 0.0%–5.6%) |
| **Any Local** | 12.4% (12, 95% CI 6.6%–20.6%) | 8.2% (8, 95% CI 3.6%–15.6%) |
| **Any Systemic** | 13.4% (13, 95% CI 7.3%–21.8%) | 12.4% (12, 95% CI 6.6%–20.6%) |
| **<7 Days** | 21.6% (21, 95% CI 13.9%–31.2%) | 18.6% (18, 95% CI 11.4%–27.7%) |
| **8-30 Days** | 4.1% (4, 95% CI 1.1%–10.2%) | 2.1% (2, 95% CI 0.3%–7.3%) |

| 18-64 years old | | |
|---|---|---|
| Event | After 1st dose<br>% (n, CI 95% Clopper-Pearson) | After 2st dose<br>% (n, CI 95% Clopper-Pearson) |
| **Local reactions** (at the application site) | | |
| Pain | 20.4% (10, 95% CI 10.2%–34.3%) | 6.1% (3, 95% CI 1.3%–16.9%) |
| Hematoma/Ecchymosis/Hypersensitivity | 2.0% (1, 95% CI 0.1%–10.9%) | - |
| Erythema/Redness | - | 2.0% (1, 95% CI 0.1%–10.9%) |
| **Systemic** | | |
| Fever | 6.1% (3, 95% CI 1.3%–16.9%) | 2.0% (1, 95% CI 0.1%–10.9%) |
| Nausea | - | - |
| Flu-like ilness | 4.1% (2, 95% CI 0.5%–14.0%) | 2.0% (1, 95% CI 0.1%–10.9%) |
| Diarrhea | 4.1% (2, 95% CI 0.5%–14.0%) | - |
| Malaise/Fatigue/Myalgia | 6.1% (3, 95% CI 1.3%–16.9%) | 4.1% (2, 95% CI 0.5%–14.0%) |
| Headache | - | 2.0% (1, 95% CI 0.1%–10.9%) |
| Retro-orbital pain | - | 2.0% (1, 95% CI 0.1%–10.9%) |
| Chest pain | - | - |
| Acute coronary syndrome | - | - |
| **Any Local** | 22.4% (11, 95% CI 11.8%–36.6%) | 8.2% (4, 95% CI 2.3%–19.6%) |
| **Any Systemic** | 20.4% (10, 95% CI 10.2%–34.3%) | 12.2% (6, 95% CI 4.6%–24.8%) |
| **<7 Days** | 24.5% (12, 95% CI 13.3%–38.9%) | 20.4% (10, 95% CI 10.2%–34.3%) |
| **8-30 Days** | 4.1% (2, 95% CI 0.5%–14.0%) | 0.0% (0, 95% CI 0.0%–7.3%) |

| 65+ years old | | |
|---|---|---|
| Event | After 1st dose<br>% (n, CI 95% Clopper-Pearson) | After 2st dose<br>% (n, CI 95% Clopper-Pearson) |
| **Local reactions** (at the application site) | | |
| Pain | 2.1% (1, 95% CI 0.1%–11.1%) | - |
| Hematoma/Ecchymosis/Hypersensitivity | - | 4.2% (2, 95% CI 0.5%–14.3%) |
| Erythema/Redness | - | 4.2% (2, 95% CI 0.5%–14.3%) |
| **Systemic** | | |
| Fever | - | 2.1% (1, 95% CI 0.1%–11.1%) |
| Nausea | 2.1% (1, 95% CI 0.1%–11.1%) | - |
| Flu-like ilness | - | 4.2% (2, 95% CI 0.5%–14.3%) |
| Diarrhea | - | - |
| Malaise/Fatigue/Myalgia | - | 2.1% (1, 95% CI 0.1%–11.1%) |
| Headache | - | 2.1% (1, 95% CI 0.1%–11.1%) |
| Retro-orbital pain | 2.1% (1, 95% CI 0.1%–11.1%) | - |
| Chest pain | 2.1% (1, 95% CI 0.1%–11.1%) | - |
| Acute coronary syndrome | - | 2.1% (1, 95% CI 0.1%–11.1%) |
| **Any Local** | 2.1% (1, 95% CI 0.1%–11.1%) | 8.3% (4, 95% CI 2.3%–20.0%) |
| **Any Systemic** | 6.3% (3, 95% CI 1.3%–17.2%) | 20.8% (10, 95% CI 10.5%–35.0%) |
| **<7 Days** | 4.2% (2, 95% CI 0.5%–14.3%) | 16.7% (8, 95% CI 7.5%–30.2%) |
| **8-30 Days** | 4.2% (2, 95% CI 0.5%–14.3%) | 4.2% (2, 95% CI 0.5%–14.3%) |

| DENV-naïve | | |
|---|---|---|
| Event | After 1st dose<br>% (n, CI 95% Clopper-Pearson) | After 2st dose<br>% (n, CI 95% Clopper-Pearson) |
| **Local reactions** (at the application site) | | |
| Pain | 16.3% (8, 95% CI 7.3%–29.7%) | 2.0% (1, 95% CI 0.1%–10.9%) |
| Hematoma/Ecchymosis/Hypersensitivity | - | 2.0% (1, 95% CI 0.1%–10.9%) |
| Erythema/Redness | - | 2.0% (1, 95% CI 0.1%–10.9%) |
| **Systemic** | | |
| Fever | 4.1% (2, 95% CI 0.5%–14.0%) | 4.1% (2, 95% CI 0.5%–14.0%) |
| Nausea | 4.1% (2, 95% CI 0.5%–14.0%) | - |
| Flu-like ilness | 2.0% (1, 95% CI 0.1%–10.9%) | 4.1% (2, 95% CI 0.5%–14.0%) |
| Diarrhea | - | - |
| Malaise/Fatigue/Myalgia | 2.0% (1, 95% CI 0.1%–10.9%) | 2.0% (1, 95% CI 0.1%–10.9%) |
| Headache | - | 2.0% (1, 95% CI 0.1%–10.9%) |
| Retro-orbital pain | - | - |
| Chest pain | - | - |
| Acute coronary syndrome | - | - |
| **Any Local** | 16.3% (8, 95% CI 7.3%–29.7%) | 6.1% (3, 95% CI 1.3%–16.9%) |
| **Any Systemic** | 12.2% (6, 95% CI 4.6%–24.8%) | 18.4% (9, 95% CI 8.8%–32.0%) |
| **<7 Days** | 24.5% (12, 95% CI 13.3%–38.9%) | 18.4% (9, 95% CI 8.8%–32.0%) |
| **8-30 Days** | 6.1% (3, 95% CI 1.3%–16.9%) | 0.0% (0, 95% CI 0.0%–7.3%) |

| DENV-exposed | | |
|---|---|---|
| Event | After 1st dose<br>% (n, CI 95% Clopper-Pearson) | After 2st dose<br>% (n, CI 95% Clopper-Pearson) |
| **Local reactions** (at the application site) | | |
| Pain | 6.3% (3, 95% CI 1.3%–17.2%) | 4.2% (2, 95% CI 0.5%–14.3%) |
| Hematoma/Ecchymosis/Hypersensitivity | 2.1% (1, 95% CI 0.1%–11.1%) | 2.1% (1, 95% CI 0.1%–11.1%) |
| Erythema/Redness | - | 4.2% (2, 95% CI 0.5%–14.3%) |
| **Systemic** | | |
| Fever | 2.1% (1, 95% CI 0.1%–11.1%) | - |
| Nausea | 2.1% (1, 95% CI 0.1%–11.1%) | - |
| Flu-like ilness | 2.1% (1, 95% CI 0.1%–11.1%) | 2.1% (1, 95% CI 0.1%–11.1%) |
| Diarrhea | 4.2% (2, 95% CI 0.5%–14.3%) | - |
| Malaise/Fatigue/Myalgia | - | 4.2% (2, 95% CI 0.5%–14.3%) |
| Headache | - | 2.1% (1, 95% CI 0.1%–11.1%) |
| Retro-orbital pain | - | 2.1% (1, 95% CI 0.1%–11.1%) |
| Chest pain | 2.1% (1, 95% CI 0.1%–11.1%) | - |
| Acute coronary syndrome | - | 2.1% (1, 95% CI 0.1%–11.1%) |
| **Any Local** | 8.3% (4, 95% CI 2.3%–20.0%) | 10.4% (5, 95% CI 3.5%–22.7%) |
| **Any Systemic** | 12.5% (6, 95% CI 4.7%–25.2%) | 12.5% (6, 95% CI 4.7%–25.2%) |
| **<7 Days** | 18.8% (9, 95% CI 8.9%–32.6%) | 18.8% (9, 95% CI 8.9%–32.6%) |
| **8-30 Days** | 2.1% (1, 95% CI 0.1%–11.1%) | 4.2% (2, 95% CI 0.5%–14.3%) |

Suppl. Table 2
