## Supplementary Table S3 for "Serotype skewing and immune imprinting shape response to the tetravalent dengue virus Qdenga vaccine"

**Geometric Mean Titer - 95% CI**

| | DENV-naive | DENV-exposed |
|---|---|---|
| **DENV 1** | | |
| T0 | 10.086 (10.035–10.138) | 95.147 (58.773-154.034) |
| T1 | 40.037 (26.672–60.099) | 752.365 (431.049-1313.199) |
| T2 | 43.758 (28.913–66.225) | 936.483 (556.819-1575.019) |
| T3 | 47.565 (30.098–75.171) | 1094.213 (629.452-1902.134) |
| **DENV 2** | | |
| T0 | 10.835 (10.115–11.607) | 24.417 (17.569-33.933) |
| T1 | 52.820 (37.876–73.660) | 180.751 (125.778-259.752) |
| T2 | 63.301 (44.390–90.267) | 181.621 (128.661-256.381) |
| T3 | 81.517 (54.538–121.843) | 170.953 (118.204-247.242) |
| **DENV 3** | | |
| T0 | 11.540 (9.389–14.186) | 25.828 (18.306-36.440) |
| T1 | 41.580 (30.415–56.843) | 194.911 (116.765-325.358) |
| T2 | 27.966 (21.123–37.026) | 141.933 (85.287-236.201) |
| T3 | 32.107 (23.902–43.129) | 232.667 (151.340-357.698) |
| **DENV 4** | | |
| T0 | 10.056 (9.997–10.115) | 10.868 (10.195-11.585) |
| T1 | 11.619 (10.258–13.160) | 19.225 (15.367-24.053) |
| T2 | 11.449 (10.133–12.936) | 18.885 (14.970-23.824) |
| T3 | 11.764 (10.510–13.168) | 17.830 (14.332-22.184) |

Suppl. Table 3
